# Unsupervised machine-learning identifies clinically distinct subtypes of ALS that reflect different genetic architectures and biological mechanisms

**DOI:** 10.1101/2023.06.12.23291304

**Authors:** Thomas P Spargo, Heather Marriott, Guy P Hunt, Oliver Pain, Renata Kabiljo, Harry Bowles, William Sproviero, Alexandra C Gillett, Isabella Fogh, Project MinE ALS Sequencing Consortium, Peter M. Andersen, Nazli A. Başak, Pamela J. Shaw, Philippe Corcia, Philippe Couratier, Mamede de Carvalho, Vivian Drory, Jonathan D. Glass, Marc Gotkine, Orla Hardiman, John E. Landers, Russell McLaughlin, Jesús S. Mora Pardina, Karen E. Morrison, Susana Pinto, Monica Povedano, Christopher E. Shaw, Vincenzo Silani, Nicola Ticozzi, Philip Van Damme, Leonard H. van den Berg, Patrick Vourc’h, Markus Weber, Jan H. Veldink, Richard J.B. Dobson, Ahmad Al Khleifat, Nicholas Cummins, Daniel Stahl, Ammar Al-Chalabi, Alfredo Iacoangeli

**Affiliations:** Maurice Wohl Clinical Neuroscience Institute, King’s College London, Department of Basic and Clinical Neuroscience, London, UK; Department of Biostatistics and Health Informatics, King’s College London, London, UK; NIHR Maudsley Biomedical Research Centre (BRC) at South London and Maudsley NHS Foundation Trust and King’s College London, London, UK; Perron Institute for Neurological and Translational Science, Nedlands, WA 6009, Australia; Centre for Molecular Medicine and Innovative Therapeutics, Murdoch University, Murdoch, WA 6150, Australia; Department of Psychiatry, University of Oxford, Oxford, UK; Social, Genetic and Developmental Psychiatry Centre, Institute of Psychiatry, Psychology and Neuroscience, King’s College London, London, UK; Department of Clinical Science, Umeå University, Umeå SE-901 85, Sweden; Koc University, School of Medicine, Translational Medicine Research Center, NDAL, Istanbul, 34010, Turkey; Sheffield Institute for Translational Neuroscience (SITraN), University of Sheffield, Sheffield S10 2HQ, UK; UMR 1253, Université de Tours, Inserm, Tours 37044, France; Centre de référence sur la SLA, CHU de Tours, Tours 37044, France; Centre de référence sur la SLA, CHRU de Limoges, Limoges, France; UMR 1094, Université de Limoges, Inserm, Limoges 87025, France; Instituto de Fisiologia, Instituto de Medicina Molecular João Lobo Antunes, Faculdade de Medicina, Universidade de Lisboa, Lisbon 1649-028, Portugal; Department of Neurology, Tel-Aviv Sourasky Medical Centre, Tel-Aviv 64239, Israel; Sackler Faculty of Medicine, Tel-Aviv University, Tel-Aviv 6997801, Israel; Department of Neurology, Emory University School of Medicine, Atlanta, Georgia, GA 30322, USA; Faculty of Medicine, Hebrew University of Jerusalem, Jerusalem 91904, Israel; Agnes Ginges Center for Human Neurogenetics, Department of Neurology, Hadassah Medical Center, Jerusalem 91120, Israel; Academic Unit of Neurology, Trinity Biomedical Sciences Institute, Trinity College Dublin, Dublin D02 PN40, Ireland; Department of Neurology, UMass Chan Medical School, Worcester, MA 01655, USA; Complex Trait Genomics Laboratory, Smurfit Institute of Genetics, Trinity College Dublin, Dublin D02 PN40, Ireland; ALS Unit, Hospital San Rafael, Madrid, Spain; School of Medicine, Dentistry and Biomedical Sciences, Queen’s University Belfast, Belfast BT9 7BL, UK; Functional Unit of Amyotrophic Lateral Sclerosis (UFELA), Service of Neurology, Bellvitge University Hospital, L’Hospitalet de Llobregat, Barcelona 08907, Spain; Department of Neurology Stroke Unit and Laboratory of Neuroscience, Istituto Auxologico Italiano, IRCCS, Milan 20149, Italy; Department of Pathophysiology and Transplantation, “Dino Ferrari” Center, Università degli Studi di Milano, Milan 20122; Department of Neurology, University Hospitals Leuven and Department of Neuroscience, KU Leuven, Leuven 3000, Belgium; VIB, Center for Brain and Disease Research, Leuven 3000, Belgium; Department of Neurology, UMC Utrecht Brain Center, University Medical Center Utrecht 3584 CX, Netherlands; Service de Biochimie et Biologie molécularie, CHU de Tours, Tours 37044, France; Neuromuscular Diseases Unit/ALS Clinic, Kantonsspital St. Gallen, 9007 St. Gallen, Switzerland; Institute of Health Informatics, Universit College London, London, UK; King’s College Hospital, Bessemer Road, London, SE5 9RS, UK

**Keywords:** amyotrophic lateral sclerosis, patient stratification, latent class analysis, machine learning, clustering

## Abstract

**Background:** Amyotrophic lateral sclerosis (ALS) is a fatal neurodegenerative disease characterised by a highly variable clinical presentation and multifaceted genetic and biological bases that translate into great patient heterogeneity. The identification of homogeneous subgroups of patients in terms of both clinical presentation and biological causes, could favour the development of effective treatments, healthcare, and clinical trials. We aimed to identify and characterise homogenous clinical subgroups of ALS, examining whether they represent underlying biological trends.

**Methods:** Latent class clustering analysis, an unsupervised machine-learning method, was used to identify homogenous subpopulations in 6,523 people with ALS from Project MinE, using widely collected ALS-related clinical variables. The clusters were validated using 7,829 independent patients from STRENGTH. We tested whether the identified subgroups were associated with biological trends in genetic variation across genes previously linked to ALS, polygenic risk scores of ALS and related neuropsychiatric traits, and in gene expression data from post-mortem motor cortex samples.

**Results:** We identified five ALS subgroups based on patterns in clinical data which were general across international datasets. Distinct genetic trends were observed for rare variants in the *SOD1* and *C9orf72* genes, and across genes implicated in biological processes relevant to ALS. Polygenic risk scores of ALS, schizophrenia and Parkinson’s disease were also higher in distinct clusters with respect to controls. Gene expression analysis identified different altered biological processes across clusters reflecting the genetic differences. We developed a machine learning classifier based on our model to assign subgroup membership using clinical data available at first visit, and made it available on a public webserver at http://latentclusterals.er.kcl.ac.uk.

**Conclusion:** ALS subgroups characterised by highly distinct clinical presentations were discovered and validated in two large independent international datasets. Such groups were also characterised by different underlying genetic architectures and biology. Our results showed that data-driven patient stratification into more clinically and biologically homogeneous subtypes of ALS is possible and could help develop more effective and targeted approaches to the biomedical and clinical study of ALS.

## Introduction

Amyotrophic lateral sclerosis (ALS) is a devastating neurodegenerative disease characterised by progressive neuromuscular degeneration leading to death, typically from respiratory failure within three years of onset (1). The disease affects upper and lower motor neurons in the brain and spinal cord and has an estimated lifetime risk of between 1 in 300-400 people.

ALS is clinically defined, yet its clinical presentation varies greatly. The mean age of onset is around 60 years although people may develop ALS at almost any age during adulthood (2). ALS normally leads to death within 3-5 years of onset, however, in some patients it can occur within a year, and 5-10% of people live for over 10 years (3–5). Between 60 and 70% of people first develop symptoms in the spinal innervated muscles, with others having bulbar, mixed or, in about 3%, respiratory onset (2, 6–8). The extent of involvement of upper and lower motor neurons varies, ranging between a pure upper motor neuron (primary lateral sclerosis; PLS) and pure lower motor neuron phenotype (progressive muscular atrophy; PMA), with most presentations being a mixture of the two (9, 10). An overlap with frontotemporal dementia (FTD) is also recognised, with a joint diagnosis for up to 15% of people in some studies and cognitive dysfunction in around 50%, according to disease stage (11, 12).

The genetic landscape of ALS is similarly heterogenous, with recognised monogenic, oligogenic, and polygenic contributions to disease (1, 13–17). Known genetic variation explains disease for around 15-20% of people, implicating variants in over 40 genes as causal for or modifiers of ALS (15, 18, 19). Likewise, ALS is associated with disruption to various biological processes, including cytoskeletal transport, RNA function, autophagy, and proteostasis (1, 7, 20).

Unfortunately, the efficacy of existing treatments for ALS is limited; the most effective drug therapies extend life expectancy from onset by no more than a few months (21, 22). This poor efficacy is partially due to the heterogeneity of the disease, and therefore more effective treatments may be discovered within a precision medicine framework. Existing research supports this hypothesis. For instance, treatment with lithium appears to extend survival trajectories specifically among people with an *UNC13A* variant (23). Further research examining the utility of gene therapies that aim to offset aberrant gene function associated with specific genetic variation is ongoing (24). This is valuable for *SOD1*-ALS which appears biologically separate from non-*SOD1* ALS (25) but may be suboptimal when the biological disease signature is indistinguishable between people with and without a given variant, as for *C9orf72* and non-*C9orf72* associated ALS (26), given the breadth of processes implicated in the disease.

Patient stratification is an effective avenue for discovery of biological mechanisms relevant to particular subgroups (27). Such stratification has been attempted previously. For instance, clusters of ALS have been identified based on biological trends in transcriptomic and neuroanatomical data, with evidence suggesting that these groups may be reflected in the phenotype (28–33).

Previous attempts to clustering in ALS have been limited to individual national cohorts and were not generalisable. For example, clinical clusters, predictive of disease duration, were identified in a British cohort using latent class cluster analysis (LCA) (34). Independently, semi-supervised machine-learning applied to high-dimensional clinical data from two independent Italian cohorts identified clusters conforming to the following clinical subgroups: bulbar, respiratory, flail arm, classical, pyramidal, and flail leg (35).

No study to date has validated the subgroups they identified using independent samples from different populations which greatly limits their applicability. Moreover, despite ALS being clinically defined, no study has examined whether data-driven clinical subgroups differ biologically beyond subgroups defined by individual gene variants. The identification of clinically and biologically homogenous subgroups that are robust and consistent across populations of patients will likely benefit development of precision medicine approaches for ALS extending beyond those targeting specific genetic vulnerabilities.

Accordingly we examined: (A) whether data-driven clusters defined by widely collected clinical measures could be identified and validated across international ALS cohorts; (B) clinical characteristics defining these clusters; (C) whether clusters differ biologically in terms of (i) the frequency of rare variants in genes previously associated with ALS, (ii) common genetic variation captured within polygenic risk scores (PRS) for risk of ALS and related neuropsychiatric disorders, and (iii) molecular signatures via gene expression levels; (D) the extent to which clinically driven clusters could be identified using data attainable around the time of diagnosis. Across these investigations we used data of over 14,000 ALS patients from the Project MinE (36), the Survival, Trigger and Risk, Epigenetic, eNvironmental and Genetic Targets for motor neuron Health (STRENGTH) consortia, and the King’s College London (KCL) Brain Bank (37, 38).

## Methods

### Sample

People with a diagnosis of ALS, PMA, or PLS were sampled from two international consortia: Project MinE (36) and STRENGTH. All samples underwent pre-processing (see Figure 1) to determine the sample entered into LCA and subsequent analyses (N: Project MinE = 6,523, STRENGTH = 7,829). Missingness was present in both cohorts (see Figure S1). This is handled in LCA using a full information maximum likelihood approach, which enables model fitting for records with incomplete data (39). Subsequent analyses employed a complete case analysis approach, retaining only people with information recorded for all variables used in that analysis.

**Figure 1.**
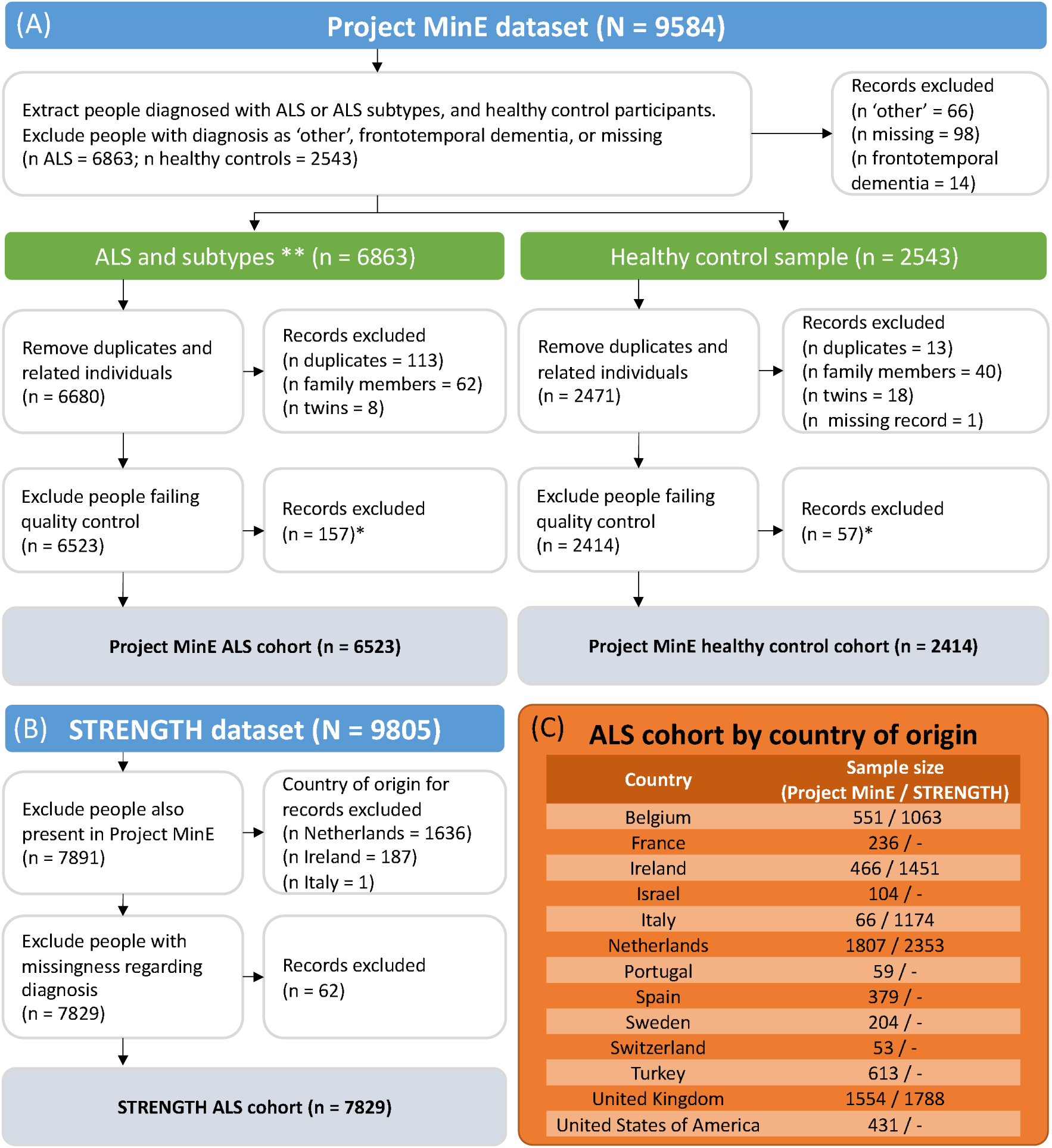
Summary of data processing and samples available by country for the Project MinE and STRENGTH cohorts. ALS = amyotrophic lateral sclerosis, PLS = primary lateral sclerosis, PMA = progressive muscular atrophy. *Quality control procedures for Project MinE have been described previously (cf. (14, 36, 40)); **ALS clinical diagnosis encompasses recorded as: ALS, ALS/frontotemporal dementia, ALS/PLS, ALS/PMA, or progressive bulbar palsy (cf. (9)); PLS and PMA are retained as distinct ALS subtypes in all analysis. **Panel A:** processing for people with ALS and healthy controls from the whole-genome sequencing cohort from Project MinE; **Panel B:** processing of the clinical ALS dataset from STRENGTH; **Panel C:** distribution of people in the final ALS cohort across countries.

Age- and sex-matched control participants from Project MinE (N = 2,414 after pre-processing; see Figure 1) were also sampled as a comparison group for analysis of trends in common genetic variation.

### Study design

#### Clinical data

Phenotypic information from the Project MinE and STRENGTH datasets were used as features for clustering. The included variables are all frequently collected for people with ALS: sex at birth (male or female), site of onset (not-bulbar or bulbar), clinical diagnosis (ALS, PLS, or PMA), age of onset (years), disease duration (years) from onset until death or last status update with associated censoring status (alive or deceased), and delay from onset until diagnosis. Diagnostic delay was standardised by country to account for any inter-country differences (see Figure S2). Standardisation was performed by centring each person’s diagnostic delay by the per-country mean and scaling relative to the per-country standard deviation (see Table S1).

#### Genetic data

Associations between clinically-defined clusters and biological trends were performed using data from Project MinE. Whole-genome sequence data were generated as previously described (36).

Information on rare genetic variation was extracted for a panel of 36 genes previously implicated in ALS: *ALS2, ANG, ANXA11, ATXN1, ATXN2, CFAP410* (formerly *C21orf2*)*, CHCHD10, CHMP2B, C9orf72, DAO, DCTN1, ERBB4, FIG4, FUS, hnRNPA1, MATR3, MOBP, NEFH, NEK1, OPTN, PFN1, SCFD1, SETX, SIGMAR1, SOD1, SPG11, SQSTM1 (p62), TAF15, TARDBP, TBK1, TUBA4A, UBQLN2, UNC13A, VAPB, VCP, VEGFA*. Summaries of variants occurring across Project MinE samples are available in the databrowser (40).

Variation in *C9orf72* and *ATXN2* was reported regarding presence or absence of known pathogenic short tandem repeat expansions in each gene. Repeat expansion lengths were determined using Expansion Hunter (41). *C9orf72* expansions were confirmed using repeat-primed PCR, being classified as inconsistent if the dry and wet lab results did not match (42, 43). The minimum number of repeat units denoting presence of a repeat expansion was 30 for *C9orf72* and 28 for *ATXN2* (44). For *C9orf72,* inconsistent expansions were reported in 32 people and coded as missing.

The presence or absence of rare variants (MAF <0.01 in both Project MinE controls and Gnomad controls v2.1.1) predicted by the Ensembl Variant Effect Predictor (45) to have a high or moderate impact upon gene function was recorded across the remaining 34 genes. Moderate impact variants included missense, in-frame insertions and deletions, and protein altering variants. High impact variants included stop lost and gained, start lost, transcript amplification, frameshift, transcript ablation and splice acceptor and donor variants.

Variants across all genes except *C9orf72* and *SOD1* were aggregated into burden groups (see Table S2). The main burden groups described three functional pathways related to cellular processes disrupted in ALS: ‘autophagy and proteostasis’, ‘RNA function’, and ‘cytoskeletal dynamics and axonal transport’. Genes were assigned to pathways according to the involvement of their protein products within them and to the processes which are disrupted by deleterious variation in each gene (see Table S2); this was determined through literature review. A further ‘any pathway’ group, aggregating across the three functional pathways, was also defined. Each burden group was binary-coded according to presence or absence of variants in at least one gene assigned to the group.

The *SOD1* and *C9orf72* genes were analysed individually since their variants are the most frequent genetic causes of ALS, likely reflecting that they are implicated in various disease pathways (46, 47), and each occurred with sufficient frequency to be tested individually.

Associations between clusters and common genetic variation were examined using PRS indicating risk for ALS and related neuropsychiatric diseases. PRS were derived from European ancestry genome-wide association study (GWAS) summary statistics of risk for ALS (14), FTD (48), Alzheimer’s disease (49), Parkinson’s disease (50), and schizophrenia (51). Scores were calculated with SBayesR (52) under the reference-standardised approach of *GenoPredPipe* (53) (see Supplementary Methods 1.1).

Since samples from Project MinE are included within the ALS GWAS, PRS for this trait were generated based on GWAS summary statistics that exclude meta-analysis ‘stratum 6’, which includes most people from Project MinE. To ensure no sample overlap, analyses including the ALS PRS were performed using only those who were sampled within GWAS stratum 6. All available Project MinE samples were included in analyses based on PRS for other traits.

#### Gene expression data

The KCL BrainBank expression dataset consists of post-mortem bulk RNA-sequencing samples from the Medical Research Council (MRC) London Neurodegenerative Diseases Brain Bank at KCL. Frozen human post-mortem tissue was taken from the primary motor cortex of patients and controls whose genomes were sequenced as part of Project MinE. The protocols for RNA-sequencing (37, 38) has been described previously.

### Procedure

#### 1. Clustering of ALS clinical data

Latent Class Cluster Analysis (LCA) was applied to identify data-driven subgroups in clinical variables (see Supplementary Methods 1.2.). LCA is an unsupervised machine-learning approach with various benefits: information returned enables the immediate inspection of fit quality; data across categorical, ordinal, and continuous modalities can be combined, including time-to-event variables with censoring; an in-built full information maximum likelihood approach (39) enables class (cluster) assignment for people with incomplete records.

Project MinE was used as the discovery cohort, holding back independent samples from STRENGTH for model validation. After validation, the independent Project MinE and STRENGTH samples were pooled together and LCA was repeated within the joint dataset. Consistency between the discovery and joint dataset latent class models was established by inspecting cluster assignment overlap between them. The joint-dataset model was used for subsequent analyses.

#### 2. Clinical characterisation of clusters

Characteristics of the identified clusters predicted by the features used in LCA were first examined using linear discriminant analysis. This algorithm derives linear axes that maximise separation between the classes. The associations between these axes and the predictor variables indicate which variables best distinguish the classes. Although the method performs adequately with categorical data (55, 56), linear discriminant analysis assumes that predictors are continuous and normally distributed. Multinomial logistic regression (57), which makes no such assumption, was therefore applied with stepwise feature selection to support linear discriminant analysis.

Differences in disease duration between classes were examined first via pairwise log-rank tests and second within a cox proportional-hazards model, including class and the other LCA model features as covariates.

Additional procedural information is provided in supplementary methods 1.3.

#### 3. Biological trends across clusters

Associations between the *k* clusters identified and rare variation in ALS-implicated genes were investigated using *k*x2 Fisher’s exact tests, with a significance threshold of false-discovery rate adjusted p<0.05. Odds ratios for having a variant in a given cluster vs all other clusters were also determined.

Binary logistic regression models were used to compare PRS for each cluster against all other clusters and against healthy controls. Reflecting the non-independence of these tests, significance was defined by a nominal p<0.05. Each analysis was performed with a given PRS as a univariate predictor and after including the first five principal components of ancestry as covariates.

To determine whether the clusters displayed alterations in biological processes, we performed differential expression and gene enrichment analysis for the samples from Project MinE which had matching motor cortex expression data available (N = 88) and 59 controls. The processing pipeline used for the expression analysis is documented and available on GitHub (https://github.com/rkabiljo/RNASeq_Genes_ERVs and https://github.com/rkabiljo/DifferentialExpression_Genes). Briefly, reads are first interleaved using reformat.sh from BBtools (58), adapters and low-quality reads are trimmed using bbduk.sh (58). The data is then aligned to Hg38 using STAR (59). Transcripts are quantified using HTSeq (60). DESeq2 (61) is used for normalisation and differential expression analysis. An extensive description of the pipeline was recently published (62). Gene enrichment analysis was performed using the most significant 500 differentially expressed genes with clusterProfiler (63).

Further procedural details are provided in supplementary methods 1.4.

#### 4. Prediction of cluster membership using baseline data

Random forest and eXtreme Gradient Boosting classification algorithms were trained to examine whether cluster membership could be predicted using only information accessible around the time of first diagnosis. Six algorithms were trained across the two machine-learning methods with a multiclass classification objective, predicting assignments to the LCA-identified clusters, across three data configurations: first, all clinical features used in LCA except disease duration (which cannot be assessed at this time); second, clinical features from model 1 alongside rare and common genetic variables used to assess biological trends across classes; third, clinical features from model 1, sample-matched with model 2.

Shapley Additive exPlanations (SHAP) (64, 65) were used to examine which features were more influential upon machine-learning algorithm class membership predictions. To further examine which features were important for prediction of exclusively Class 1 versus 2, the largest LCA clusters, 6 additional binary classification algorithms were trained across the two machine-learning methods and three data configurations after restricting only to people assigned to these classes.

Only people without missingness in included features were used in each multiclass (binary) analysis, therefore the total sample size was 12,508 (11,109) in the first data configuration and 3,226 (2,996) in the second and third. The Algorithms were trained with 10-fold cross-validation, repeated 10 times using pseudorandom seeding. Out-of-fold prediction performance was evaluated using the metrics of sensitivity, specificity, precision, and balanced accuracy.

## Results

### 1. Clustering of ALS clinical data

We identified that clinical subgroups of ALS were well described within a 5-class latent class model (see Table S3; Table 1).

**Table 1.** Class membership characteristics for a 5-class model in the Project MinE, STRENGTH, and joint datasets. Numbers presented in bold refer to average posterior probability of belonging to the assigned class. The statistics in the discovery and validation datasets are for the 5-class model fitted to the discovery sample. The Joint dataset statistics are for the 5-class model fitted to the joint dataset which combines Project MinE and STRENGTH.

| Dataset | Assigned class | N in class (% of dataset) based on |  | Average posterior probability of belonging to class |  |  |  |  |
| --- | --- | --- | --- | --- | --- | --- | --- | --- |
|  |  | posterior probabilities | most likely class membership | 1 | 2 | 3 | 4 | 5 |
| Discovery (Project MinE) | 1 | 3702.34 (0.568) | 3952 (0.606) | **0.883** | 0.106 | 0.01 | 0.001 | 0 |
|  | 2 | 2110.49 (0.324) | 2023 (0.310) | 0.104 | **0.818** | 0.056 | 0.011 | 0.011 |
|  | 3 | 527.84 (0.081) | 409 (0.063) | 0.01 | 0.075 | **0.909** | 0.006 | 0 |
|  | 4 | 114.15 (0.018) | 87 (0.013) | 0 | 0.006 | 0.047 | **0.944** | 0.003 |
|  | 5 | 68.18 (0.010) | 52 (0.008) | 0.002 | 0.118 | 0.016 | 0.036 | **0.829** |
| Validation (STRENGTH) | 1 | 3887.36 (0.497) | 4126 (0.527) | **0.89** | 0.101 | 0.008 | 0.001 | 0 |
|  | 2 | 2606.34 (0.333) | 2452 (0.313) | 0.084 | **0.866** | 0.044 | 0.004 | 0.002 |
|  | 3 | 1009.62 (0.130) | 943 (0.120) | 0.009 | 0.07 | **0.912** | 0.008 | 0 |
|  | 4 | 263.43 (0.034) | 252 (0.032) | 0 | 0.001 | 0.041 | **0.955** | 0.003 |
|  | 5 | 62.24 (0.008) | 56 (0.007) | 0.001 | 0.006 | 0 | 0.008 | **0.984** |
| Joint | 1 | 7027.66 (0.490) | 7401 (0.516) | **0.9** | 0.093 | 0.006 | 0.001 | 0 |
|  | 2 | 5602.10 (0.390) | 5470 (0.381) | 0.067 | **0.879** | 0.044 | 0.006 | 0.004 |
|  | 3 | 1319.10 (0.092) | 1138 (0.079) | 0.003 | 0.085 | **0.898** | 0.013 | 0.001 |
|  | 4 | 302.60 (0.021) | 259 (0.018) | 0 | 0 | 0.04 | **0.957** | 0.003 |
|  | 5 | 100.54 (0.007) | 84 (0.006) | 0 | 0.067 | 0.024 | 0.019 | **0.889** |

The 5-class model was first identified in the Project MinE discovery sample, with lower Akaike information criterion (AIC) and Bayesian Information Criterion (BIC) values than models with fewer classes (Table S3). Latent class models did not converge when considering a higher number of classes. Acceptance of the 5-class model was supported by its entropy (0.791) indicating that the model had reasonable certainty in classification, and by people having a high probability of belonging to their assigned classes (Table 1). An equivalent model, with high entropy (0.850), was identified when repeating LCA after restricting to only people without missingness in diagnostic delay and disease duration (see Table S4).

The external validity of this solution was affirmed through application to independent samples from STRENGTH. High entropy (0.830) and high average class probability of belonging to assigned classes (Table 1) indicated that the solution fit these data well. Accurate prediction of class membership for people from STRENGTH within a k-nearest neighbours algorithm trained upon the clinical data from Project MinE samples further supported the validity of the subgroups identified (see Table S5).

Since the initial 5-class model fitted both the discovery and validation datasets well, LCA was repeated using a joint dataset, pooling across independent samples from Project MinE and STRENGTH. Models of 1 to 9 classes were fitted and the 5-class model was accepted above those with additional classes since the external validity of a 5-class model had already been demonstrated and because improvements to AIC and BIC were not substantial with additional classes (Table S3). Entropy in the 5-class solution remained high (0.836) and people had high probability of belonging to their assigned classes (Table 1). As before, a highly comparable model, with 0.881 entropy, was identified when repeating LCA after restricting to only people with diagnostic delay and disease duration reported (see Table S4).

Equivalence between the 5-class models fitted to the Project MinE and joint datasets was determined by examining cluster similarity: ∼91% of people from Project MinE and ∼92% of STRENGTH were assigned to the equivalent cluster in the joint dataset model (see Figure S4).

Accordingly, the joint dataset 5-class solution was accepted as the final model and used for subsequent analyses.

### 2. Clinical characterisation of clusters

We examined the clinical characteristics of the clinically-defined ALS subgroups identified with LCA. Table 2 and Figure 2 present descriptive statistics for clinical features across each class. Linear discriminant and multinomial logistic regression analyses highlight that diagnostic delay and disease duration were the main class delineators (See Figure 3; Table 3; Table S7-S8). Interestingly, although both linear discriminant 1 and 2 (LD1 and LD2) presented high correlations with diagnostic delay and disease duration, LD1 positively correlated with these clinical measures, while LD2 positively correlated with disease duration and negatively correlated with diagnostic delay (Table 3). All features were retained in the multinomial logistic regression model analysing only people without censored disease duration; sex was dropped as a predictor when people with censored disease duration were included in the analysis (Table S7-S8).

**Figure 2.**
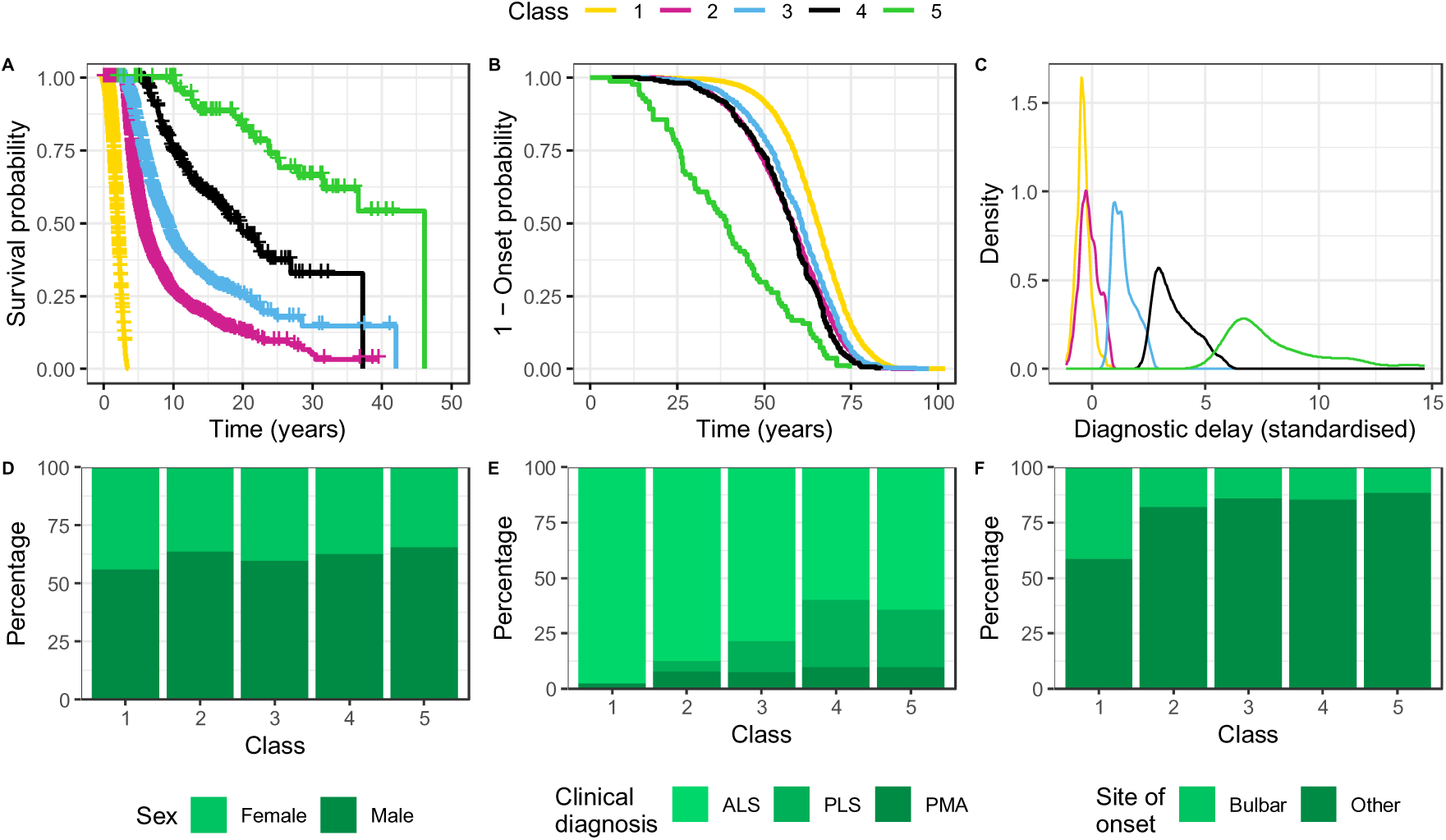
Trends in clinical features used in latent class analysis according to class. **Panel A:** Kaplan-Meier curves for disease duration from onset until death or censoring; pairwise log-rank tests indicate that survival differs significantly between all classes (p <1×10^-6^ for all comparisons after false discovery rate adjustment for all pairwise tests performed). Orthogonal tick marks on the survival curves indicate censoring. Colouring around the curves indicates 95% confidence intervals. **Panel B:** Kaplan-Meier curves for age of onset. **Panel C:** density curves for diagnostic delay centred and scaled on the mean and standard deviation for diagnostic delay according to country of origin (see Figure S2; Table S1). **Panels D-F:** stacked bar-charts indicating distribution of the categorical variables sex (D), clinical diagnosis (E), and site of onset (F) across classes. ALS = amyotrophic lateral sclerosis; PLS = primary lateral sclerosis; PMA = progressive muscular atrophy.

**Figure 3.**
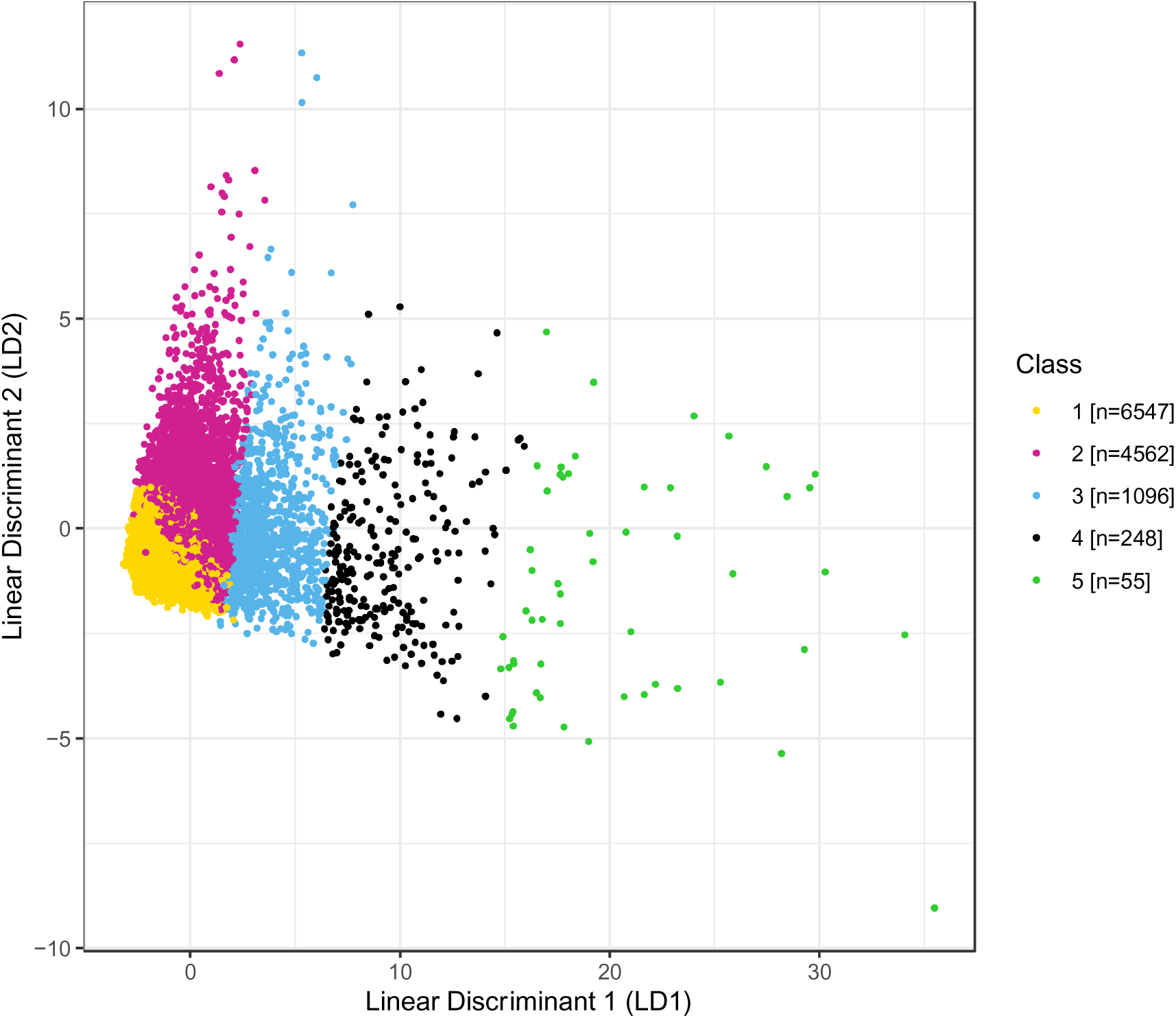
Distribution of people across the first two axes of linear discriminant analysis for all case-complete data. LD1 is highly correlated with diagnostic delay, while LD2 is associated primarily with disease duration (see Table 3). Figure S5 presents a comparison figure for LDA when restricting to only people with non-censored disease duration. Figure S6 shows this figure with people stratified by country of origin.

**Table 2.**
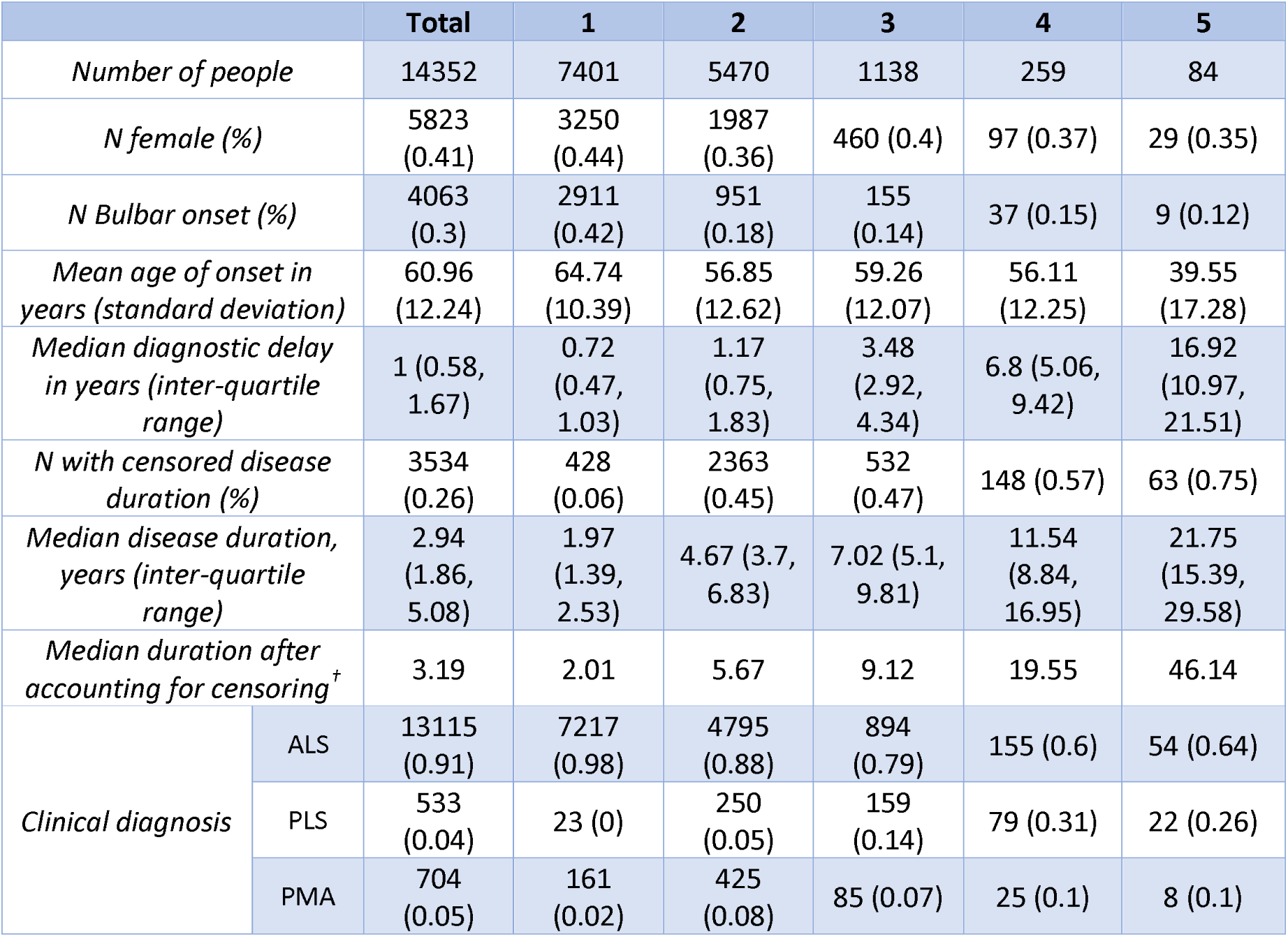
Descriptive statistics for the clinical characteristics of people with ALS across the 5-class solution fitted to the joint dataset.^†^ calculated within the survfit function of the R survival package (66); see Figure 2 for Kaplan-Meier curves stratified by class. Patterns of missingness across each variable are shown in Figure S1.

**Table 3.**
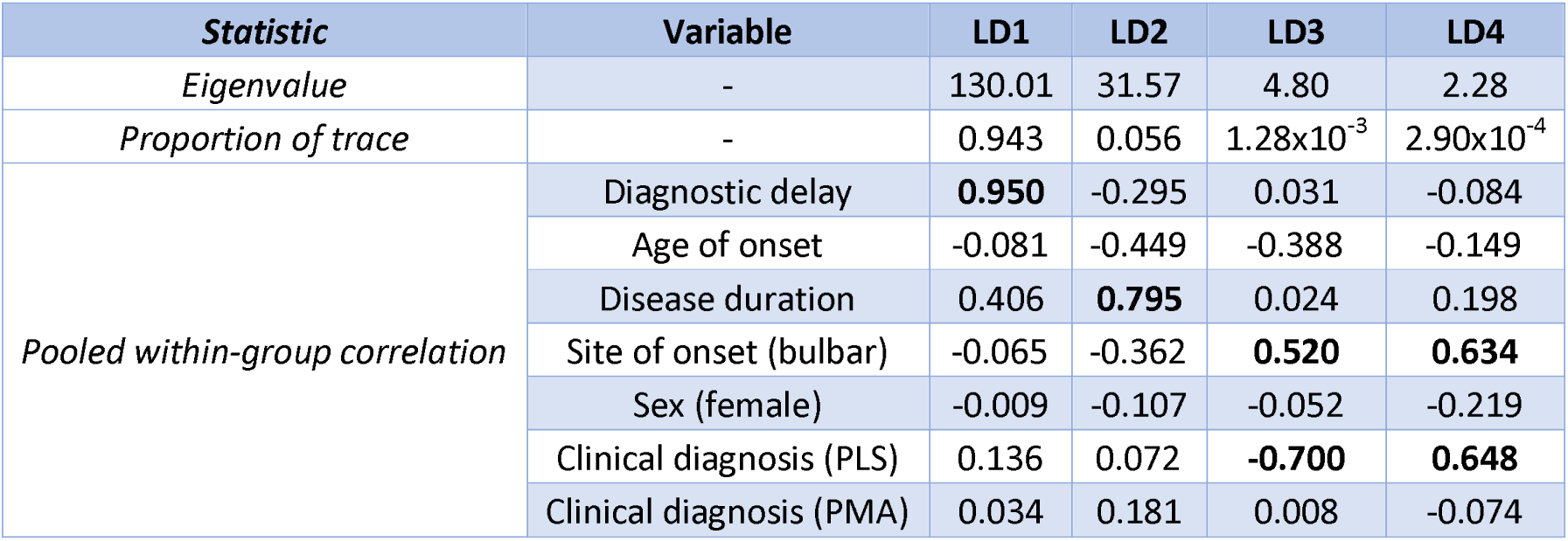
Results of the linear discriminant analysis of all people with no missingness across clinical features used in latent class analysis (n=12,508). The proportion of trace describes the proportion of the separation between classes accounted for by each linear discriminant (LD) axis. Pooled within-group correlations greater than 0.5 are presented in bold and are considered variables associated with a given LD. Reference groups for categorical variables are: ‘not-bulbar’ for site of onset, ‘male’ for sex, ‘amyotrophic lateral sclerosis’ for clinical diagnosis. This analysis includes people with censored disease duration; Table S6 presents comparable results for this analysis after restricting to only non-censored individuals. Figure 3 visualises the distribution of people and classes across the first two LD axes. PLS = primary lateral sclerosis; PMA = progressive muscular atrophy.

Survival analysis further demonstrated the relationship between class and disease duration, indicating that the classes are each associated with distinct survival trajectories (Figure 2), and class remains the most influential predictor of survival within a cox proportional-hazards model after adjusting for other clinical features (Table S9).

### 3. Biological trends across clusters

Fisher’s exact tests were applied to examine associations between rare genetic variation and class for variants occurring within genes previously associated with ALS (see Table 4). *SOD1* variants and the *C9orf72* expansion differed in frequency across classes. *C9orf72* expansions were overrepresented in Class 1 and underrepresented in Class 2; the opposite was observed for *SOD1* variants. The frequency of variants in genes linked to RNA processing and to cytoskeletal dynamics and axonal transport also differed across classes.

**Table 4.** Association between class and rare genetic variation in ALS-associated genes. Odds ratios are for having a variant in each class relative to all other classes; those with 95% confidence intervals (CI) which do not cross the null value of 1 are presented in bold. ^†^ False Discovery Rate (FDR) adjusted p-values <0.05 after adjusting for all Fisher’s exact tests in the column are presented in bold. ^Δ^ Sample size differs for C9orf72 comparison, n in class: 1 = 3,374, 2 = 2,494, 3 = 322, 4 = 65, 5 = 36. ^§^Any pathway refers to having a variant in any of the autophagy and proteostasis, RNA function, cytoskeletal dynamics and axonal transport pathways.

| Genetic variation<br>($P_{FDR}^{\dagger}$) | | Class (sample size) | | | | |
| --- | --- | --- | --- | --- | --- | --- |
|  |  | 1 (3,401) | 2 (2,512) | 3 (327) | 4 (65) | 5 (36) |
| *SOD1*<br>(**$2.04 \times 10^{-4}$**) | N with variant (freq) | 13<br>($3.84 \times 10^{-3}$) | 31 (0.01) | 6 (0.02) | 2 (0.03) | 1 (0.03) |
|  | Odds ratio [95% CI] | **0.28 [0.14, 0.53]** | **2.16 [1.21, 3.93]** | 2.37 [0.82, 5.62] | 3.87 [0.45, 15.31] | 3.43 [0.08, 21.27] |
| *C9orf72* repeat expansion <sup>Δ</sup><br>(**$2.42 \times 10^{-6}$**) | N with variant (freq) | 250 (0.07) | 101 (0.04) | 14 (0.04) | 1 (0.02) | 0 (0) |
|  | Odds ratio [95% CI] | 1.93 [1.53, 2.44] | 0.56 [0.44, 0.71] | 0.73 [0.39, 1.25] | 0.25 [0.01, 1.46] | 0 [0, 1.75] |
| Autophagy and proteostasis<br>(**0.546**) | N with variant (freq) | 309 (0.10) | 241 (0.11) | 24 (0.08) | 6 (0.10) | 5 (0.16) |
|  | Odds ratio [95% CI] | 0.96 [0.81, 1.15] | 1.08 [0.9, 1.28] | 0.77 [0.48, 1.18] | 1 [0.35, 2.32] | 1.59 [0.48, 4.15] |
| RNA function<br>(**0.036**) | N with variant (freq) | 319 (0.10) | 244 (0.11) | 19 (0.06) | 8 (0.14) | 0 (0) |
|  | Odds ratio [95% CI] | 1.02 [0.86, 1.21] | 1.08 [0.91, 1.29] | **0.59 [0.35, 0.94]** | 1.37 [0.56, 2.91] | 0 [0, 1.05] |
| Cytoskeletal dynamics and axonal transport<br>(**0.036**) | N with variant (freq) | 461 (0.16) | 416 (0.20) | 45 (0.16) | 9 (0.16) | 4 (0.13) |
|  | Odds ratio [95% CI] | **0.82 [0.71, 0.94]** | **1.27 [1.1, 1.46]** | 0.92 [0.65, 1.27] | 0.93 [0.4, 1.9] | 0.72 [0.18, 2.04] |
| Any pathway <sup>§</sup><br>(**0.029**) | N with variant (freq) | 979 (0.40) | 799 (0.47) | 81 (0.33) | 23 (0.55) | 8 (0.29) |
|  | Odds ratio [95% CI] | 0.9 [0.81, 1] | **1.17 [1.05, 1.31]** | **0.77 [0.58, 0.99]** | 1.29 [0.74, 2.21] | 0.67 [0.26, 1.52] |

Binary logistic regression models were fitted to examine associations between class and PRS for ALS and related neuropsychiatric disorders (see Figure 4). Class 1 was significantly associated with higher PRS for risk of ALS, compared both to patients in other classes and to healthy controls. Interestingly, PRSs for schizophrenia and Parkinson’s disease were lower in Class 1 compared to other classes, and were higher in Classes 2 and 5. PRS for schizophrenia were higher in most classes compared to controls, and those for Parkinson’s disease were higher for Classes 2 and 5.

**Figure 4.**
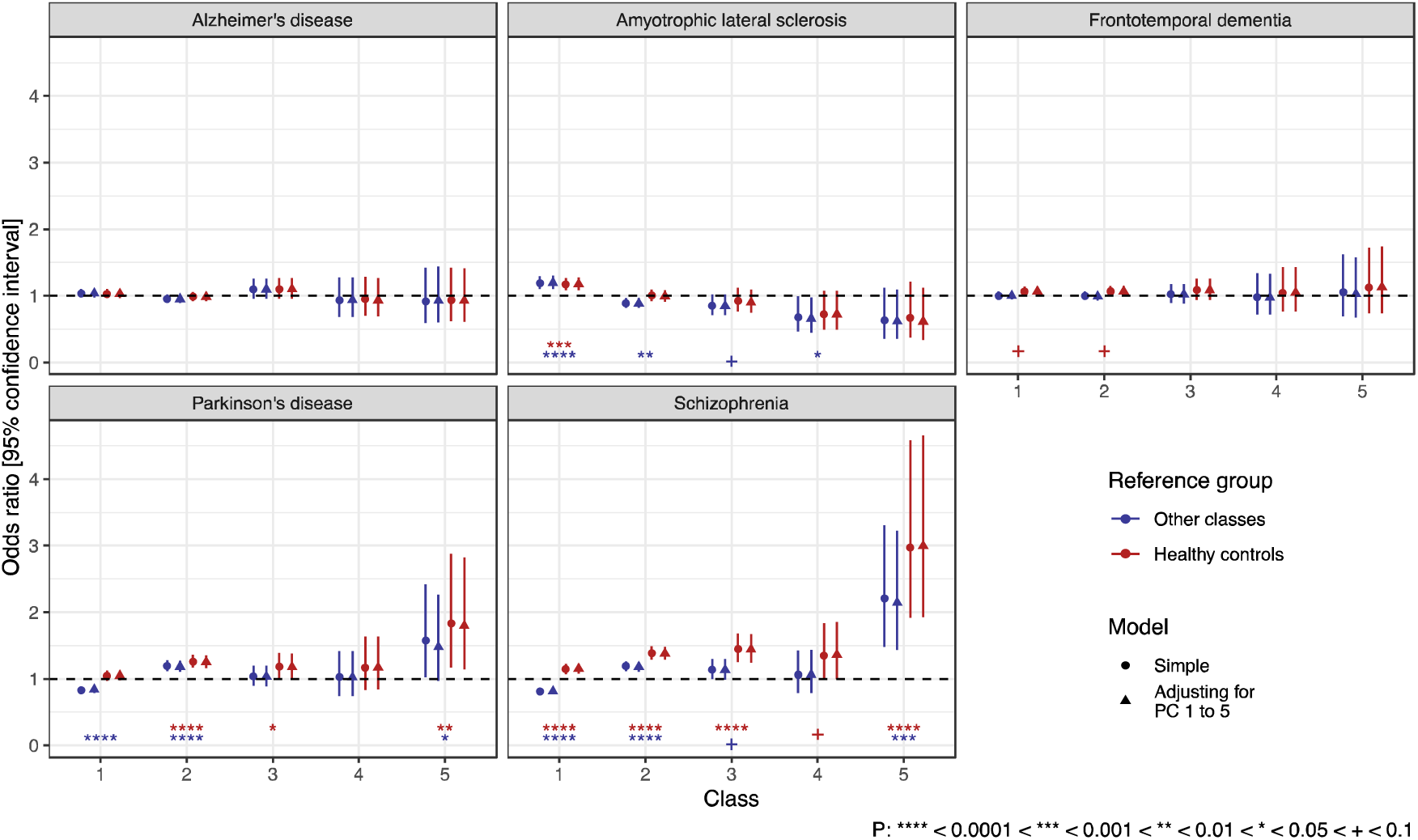
Odds ratios for association between polygenic risk scores (PRS) for neuropsychiatric diseases and class. Odds ratios are derived as exponentiated coefficients from binary logistic regression models, with the outcome variable of class vs all other classes in blue and class vs healthy controls in red. A circle denotes a ‘simple’ model including only the PRS indicated for that panel as a predictor, a triangle denotes a model which adjusts for the first five principal components (PC) of ancestry. Nominal statistical significance is indicated by asterisks for ‘simple’ models only, and colour coded according to the reference group. Sample sizes for each group in the PRS analyses [amyotrophic lateral sclerosis PRS sample size]: Class 1 = 3,464 [2,126], 2 = 2,421 [1,466], 3= 327 [203], 4 = 65 [45], 5 = 34 [20], controls = 2,371 [1,909].

RNA-sequencing data from 88 KCL BrainBank samples (a subset of Project MinE) assigned to classes via LCA (n: Class 1 = 70, Class 2 = 18) and KCL BrainBank controls (n = 59) was used to perform gene enrichment analysis of the 500 most significant differentially expressed genes present in three designs (Class 1 vs Controls, Class 1 vs Class 2, and Class 2 vs Controls). When comparing Classes 1 and 2, we found that the 500 most significant differentially expressed genes were enriched for cytoskeletal, extracellular matrix, calcium and neurotransmitter-specific synaptic signalling and muscle-function related processes, whilst both classes shared enrichments for oxidative phosphorylation and postsynaptic activity-based pathways (Figure 5). Comparisons of each class with controls highlighted processes implicated exclusively in ALS; Class 1 was enriched for ubiquitin and unfolded protein binding and nucleocytoplasmic transport, whereas Class 2 displayed enrichment for multiple neurodegenerative diseases in addition to Golgi-ER transport and T-cell signalling (Figure 5). Interestingly, these analyses showed clear reflections of the genetic differences that we found among clusters. For example, gene expression highlighted cytoskeletal and tubulin processes in the differentiation of Class 1 and 2 and so did the occurrence of rare variants in patients from these two classes. Furthermore, PRS for Parkinson’s disease was higher in Class 2 and differential expression analysis for Class 2 versus controls showed enrichment for genes involved in Parkinson’s disease pathways (KEGG). The full results are available in Tables SX1-3.

**Figure 5.**
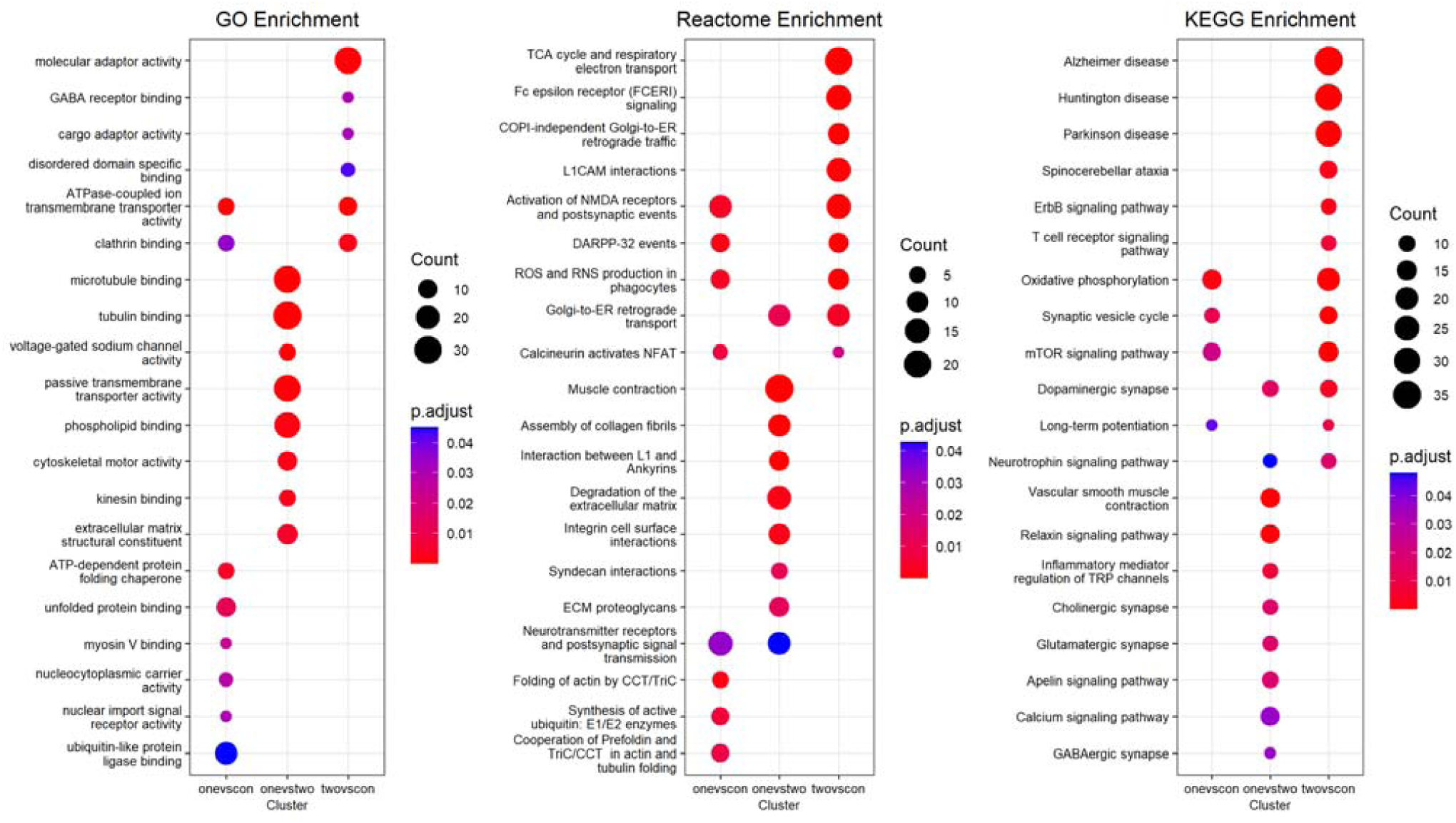
Selected results of gene enrichment analysis of the top 500 differentially expressed genes of KCL BrainBank cases assigned to classes via LDA (Class 1: n = 70, Class 2: n = 18) and controls (n = 59) in three scenarios (Class 1 vs control (onevscon), Class 1 vs Class 2 (onevstwo) and Class 2 vs Control (twovscon)). Enrichments were generated using ClusterProfiler for three databases – GO (left), Reactome (middle) and KEGG (right). Circles represent enrichments for that term with the particular scenario, with the colours denoting the p-adjusted value of the analysis. The size of the circles corresponds to the number of genes in each enrichment gene set which were present in each group. GO: Gene Ontology, KEGG: Kyoto Encyclopaedia of Genes and Genomes. The full results are available in Tables SX1-3.

### 4. Prediction of cluster membership using baseline data

We examined the extent to which machine-learning algorithms could predict class membership using data available around the time of diagnosis. Random Forest and eXtreme Gradient Boosting algorithms were trained using clinical data only or a combination of clinical and genetic variables. Comparison of the area under the receiver operating characteristic curve for prediction of each class versus all other classes determined that the two approaches performed comparably (see Figures S7-S9). Table 5 presents performance metrics for the multiclass eXtreme Gradient Boosting algorithms trained upon each data configuration. Class membership was predicted with high accuracy for Classes 3-5 (see Table 5; Figure 6). Prediction of Classes 1 and 2 still performed reasonably, but with poorer specificity in Class 1 and sensitivity in Class 2; most misclassification in these groups was for people in the opposing class (see Figures S7-S9). The algorithm performance was comparable across sample-matched datasets when using clinical features alone and using clinical and genetic measures.

**Figure 6.**
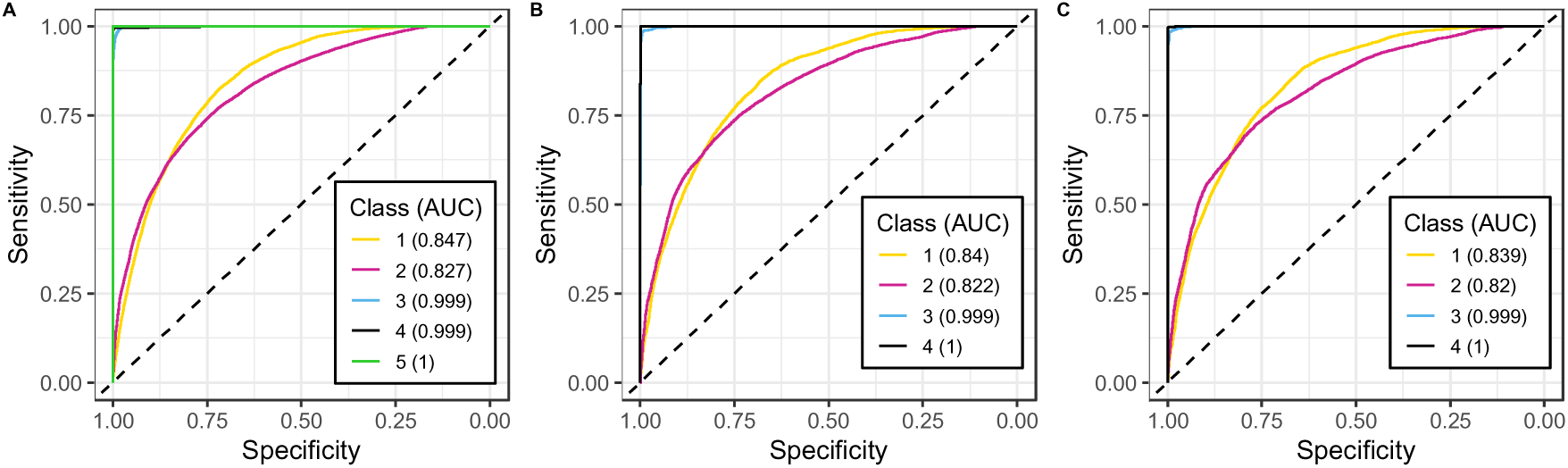
Receiver operator characteristic curves for performance of eXtreme Gradient Boosting algorithms in classifying each class versus all other classes averaged across all repeated cross-validation iterations. Each panel represents algorithms trained with different data configurations: A is upon clinical features available at diagnosis across all samples with complete clinical data (n=12,508), B is these clinical features and measured genetic risks (n=3,226), C is upon clinical features only, as in panel A, with a restricted sample to match the dataset of B (n=3,226). Class 5 is excluded from classification in panels B and C owing to small sample size in these datasets. AUC = area under the curve.

**Table 5.** Performance of eXtreme Gradient Boosting classification algorithms for predicting class membership. Class 5 was omitted from the algorithm when fewer than 20 people in the class remained in the dataset. Figures S7-S9 present receiver operating-characteristic curves and corresponding area under the curve (AUC) for each class combination pairwise.^†^ AUC values presented are for prediction of class vs all other classes (see Figure 6). *The ‘first visit clinical data [matched]’ rows describe an algorithm trained using features equivalent to the ‘first-visit clinical data’ model but after restricting the sample to match people included for the ‘First-visit clinical data and genetic features’ model.

| Model |  | Class |  |  |  |  |
| --- | --- | --- | --- | --- | --- | --- |
|  |  | 1 | 2 | 3 | 4 | 5 |
| First-visit clinical data | Sample size | 6547 | 4562 | 1096 | 248 | 55 |
|  | AUC<sup>†</sup> | 0.847 | 0.827 | 0.999 | 0.999 | 1.000 |
|  | Sensitivity | 0.77 | 0.68 | 0.97 | 0.99 | 1.00 |
|  | Specificity | 0.76 | 0.81 | 0.99 | 1.00 | 1.00 |
|  | Precision | 0.78 | 0.67 | 0.92 | 0.95 | 1.00 |
|  | Balanced Accuracy | 0.77 | 0.74 | 0.98 | 0.99 | 1.00 |
| First-visit clinical data and genetic features | Sample size | 1813 | 1177 | 192 | 44 | - |
|  | AUC<sup>†</sup> | 0.840 | 0.822 | 0.999 | 1.000 | - |
|  | Sensitivity | 0.774 | 0.685 | 0.983 | 0.977 | - |
|  | Specificity | 0.747 | 0.800 | 0.995 | 1.000 | - |
|  | Precision | 0.797 | 0.663 | 0.925 | 0.977 | - |
|  | Balanced Accuracy | 0.760 | 0.743 | 0.989 | 0.988 | - |
| First-visit clinical data [matched]<sup>*</sup> | Sample size | 1813 | 1177 | 192 | 44 | - |
|  | AUC<sup>†</sup> | 0.839 | 0.820 | 0.999 | 1.000 | - |
|  | Sensitivity | 0.776 | 0.681 | 0.984 | 0.977 | - |
|  | Specificity | 0.743 | 0.802 | 0.995 | 1.000 | - |
|  | Precision | 0.795 | 0.664 | 0.924 | 0.977 | - |
|  | Balanced Accuracy | 0.760 | 0.742 | 0.989 | 0.988 | - |

Evaluation of feature importance using SHAP values identified diagnostic delay as the most influential feature upon predictions from either approach and that various other features contributed more greatly to the prediction of Class 1 and 2 in particular (see Figure S10).

## Discussion

Latent class clustering analysis was applied to clinical data from large international ALS datasets to identify data-driven clinical subgroups and investigate whether biological differences exist between groups. Five distinct disease subgroups were identified, delineated primarily by diagnostic delay and disease duration, with importance also attributed to age and site of disease onset and clinical phenotype (ALS, PLS, or PMA). Notably, these clusters generalise across 13 countries (see Figure S6).

Survival analysis indicated that the clusters were distinct regarding disease duration; duration was greater for each respective class between 1 and 5. Diagnostic delay also generally increased with respect to class number, but, unlike disease duration, was similar in Class 1 and 2. Although diagnostic delay is commonly regarded as directly correlated with disease duration and often considered its proxy, our results suggest a more complex relationship between these two clinical measures. Indeed, the opposite signs of their correlation coefficients with LD1 and LD2 suggest the existence different patterns of progression across patients that require both diagnostic delay and disease duration to be discerned. Other features also aggregated unequally between clusters; bulbar onset was more frequent in Class 1; age of onset was later in Class 1 and earlier in Class 5, but comparable for Classes 2-4; the PLS clinical subtype was more frequent in Classes 4 and 5; the PMA subtype was more evenly distributed, but less frequent in Class 1. Figure 2 visualises the clinical characteristics of each class.

The main clinical variables discriminating subgroups from this study are consistent with those distinguishing subgroups from a previous application of LCA in a UK ALS cohort (34). Both studies find diagnostic delay as an important discriminator of ALS subgroups and that subgroups have distinct survival trajectories. However, comparing class assignments of people from STRENGTH using the latent class model from each study demonstrates that the clusters are not the same (see Figure S11). The most striking distinction is that most people assigned to Classes 1 and 2 using the present model are assigned to a single class by the model from the previous study, resulting in low resolution model that assigns the great majority of patients to only one group. This difference might be the result of two key differences between the models: the inclusion of disease duration in present model, which was withheld in the previous study and instead used as a tool for clinical validation, and the use of larger international ALS cohorts, instead of data from only one country, to train our model, which underpins its generalisability to all ALS patients.

Interestingly, the identified subgroups do not correspond to clinically defined ALS subtypes (e.g., PMA or PLS). Although this might support the hypothesis that the validity of current clinically defined subtypes should be reconsidered, it may also reflect a high rate of misdiagnosis or that the variables required to distinguish between clinical subtypes were not captured within these models. The use of high-dimensional clinical data identified ALS subgroups congruent with the classification system in a recent study (35). However, these subgroups would have fallen within the pure ALS diagnosis in our study and extreme subtypes such as PMA and PLS were not represented.

Analysis of rare genetic variants showed that the pathogenic *C9orf72* repeat expansion was more frequent in Class 1, consistent with evidence that the variant is associated with shorter disease duration (14, 67–69). Putatively deleterious *SOD1* variants were overrepresented in Class 2, the characteristics of which were more representative of the disease phenotype associated with variants in this gene than for than Class 1 (70).

Variants in genes associated with cytoskeletal dynamics and axonal transport cell processes were more frequent in Class 2. Variants in genes associated with RNA function were also unequally distributed across groups, although no individual group appeared to drive the association. These findings suggest that the ALS phenotype may present differently according to disruption of certain cellular processes. Recent studies support this possibility, reporting differences in disease progression and survival according to common variants associated with antioxidant and inflammatory disease pathways (71) and according to expression-based clusters (31, 33). Associations between disruption to particular biological processes and the ALS phenotype warrant further investigation.

Trends in common genetic variation were examined using PRS for risk of ALS or related neuropsychiatric traits. Class 1 was associated with higher ALS PRS. Conversely, Classes 2 and 5 were associated with higher schizophrenia PRS, which was also generally higher across ALS subgroups compared to healthy controls. Classes 2 and 5 were also associated with higher, and Class 1 with lower, Parkinson’s disease PRS. No associations emerged between class and PRS for FTD or Alzheimer’s disease. The lack of association with FTD PRS may reflect the limited sample size of the only available GWAS for this trait (48), and thus an underpowered PRS.

The finding that ALS PRS were only higher than controls in Class 1, the largest cluster (∼49% of all ALS), suggests that the GWAS primarily captures variants relevant to disease risk in this subgroup. Future ALS GWASs could benefit from prioritizing patients from this class to maximise their statistical power. Different variants may therefore be relevant for disease risk in the other subgroups. The possibility is supported by the association between Classes 2 and 5 and higher Parkinson’s disease PRS. These associations also suggest that genetic overlaps between ALS and other traits could be driven by certain disease subgroups (13, 14). Interestingly, the schizophrenia PRS was consistently higher than controls across all classes, supporting the hypothesis that common mechanisms are shared by all ALS patients and that these overlap at least partially with those underlying the development of schizophrenia.

Analysis of matching post-mortem motor cortex transcriptomic data available for a subset of Project MinE supported the differences in biological trends between Classes 1 and 2 in several ways. Firstly, gene enrichment analysis identified many significant processes and pathways linked to cytoskeletal dynamics and axonal transport, congruent with our finding that variants in genes linked to this pathway were more frequent in Class 2. Secondly, Class 2 was uniquely enriched for genes involved in Parkinson’s disease and schizophrenia (ErbB signalling (72, 73)), which supports our finding that Class 2 displays higher polygenic risk scores of these diseases with respect to both controls and other classes. Finding differences in oxidative stress and apoptotic processes between Classes 1 and 2 aligns well with our finding that variants in the *SOD1* gene (which codes for an antioxidant enzyme that has been proposed to affect ALS via both gain and loss of function mechanisms (47, 74)) were more frequent in Class 2. The variability in biological trends observed across these clinical subgroups supports the perspective that patient stratification may be important for identifying biological disease mechanisms (27).

Lastly, we found that class membership could be predicted with only information attainable around the time of diagnosis using machine-learning classification algorithms (see Table 5; Figure 6). Classes 3 to 5 were clearly distinguished (AUC = 0.999) and lower performance for Classes 1 and 2 (AUC_class1_ = 0.847 and AUC_class2_ = 0.827) is expected since their main delineator is disease duration which was excluded as a feature (see Table 2; Figure 2; Figure 3). Algorithms making predictions using both genetic and clinical features performed comparably to those trained using clinical features only. For rare variants in particular, this may reflect that the presence of a given variant only informs predictions for a small proportion of the whole cohort.

A limitation of this study is that rare variant analysis was conducted under the assumption that identified variants are relevant for the risk or modification of the ALS phenotype. We included rare variants predicted to have a functional effect upon genes previously implicated in ALS. Such broad inclusion criteria will likely identify a range of variants with a spectrum of relevance to the disease. Without further supporting evidence, or larger sample sizes, it is difficult to ascertain the role of each individual variant (75).

A further limitation is that biological trends could only be tested to a limited degree. Although the genetic architecture of all classes was investigated, analysis with transcriptomic data was constrained by the availability of data for relatively limited samples from only Classes 1 and 2. The genetic analyses were similarly limited by the small sample size for certain clusters. However, our investigation drew upon one of the richest genomic resources for ALS currently available (36) and this constraint should lessen in future studies as resources continue to expand.

Future studies should refine the disease classifications we have developed. The identified subgroups were partly and differentially separated by diagnostic delay and disease duration. Progression has been identified as an indicator of disease duration in ALS (76, 77), and the present patterns suggest non-linearity of progression across the disease course, which has been recognised previously (78). Our measurement of diagnostic delay is likely an imperfect proxy for disease progression. Therefore, future studies should include measures of patterns in disease progression. Recognising overlaps between ALS and other conditions (11, 14), it is also pertinent to measure non-motor features of ALS, such as cognitive or behavioural change.

In conclusion, we illustrated that our data-driven approach can identify distinct clinical subtypes of ALS and suggests that differences between these subgroups reflect the role of distinct biological mechanisms, beyond individual gene variants, upon the phenotype. The study supports the perspective that data-driven patient stratification may aid identification of biological disease mechanisms and, therefore, that such approaches should be considered in the design of future ALS studies. Defining clinically and biologically meaningful subtypes of ALS has important implications for future research and clinical practice regarding: the inherent utility of an early and reliable prediction of disease prognosis for improving and personalising patient care; improved matching of people across clinical trial placebo and active arms to facilitate testing of treatment efficacy; precision medicine development owing to easier identification of disease processes relevant for particular subgroups; the design of genetic and biological studies.

Future research should aim to develop detailed understanding of ALS subtypes by employing multi-omics datasets to examine how they are reflected across the spectrum of genome to phenome.

## Data availability

The Project Mine clinical and genetic data is available upon request at https://www.projectmine.com/research/data-sharing/. Accessing the STRENGTH clinical data requires a collaboration and data sharing agreement with the EU JPND STRENGTH consortium, administered through King’s College London (contact).

## Data Availability

All data produced in the present study are available upon reasonable request to the authors

## Acknowledgments

Samples used in this research were in part obtained from the UK National DNA Bank for MND Research, funded by the MND Association and the Wellcome Trust. We thank people with MND and their families for their participation in this project. We acknowledge sample management undertaken by Biobanking Solutions funded by the Medical Research Council at the Centre for Integrated Genomic Medical Research, University of Manchester. The authors acknowledge use of the research computing facility at King’s College London, *Rosalind* (https://rosalind.kcl.ac.uk), which is delivered in partnership with the National Institute for Health Research (NIHR) Biomedical Research Centres at South London and Maudsley and Guy’s and St. Thomas’ NHS Foundation Trusts, and part-funded by capital equipment grants from the Maudsley Charity (award 980) and Guy’s and St. Thomas’ Charity (TR130505). The authors also acknowledge the use of the CREATE research computing facility at King’s College London (79). We also acknowledge Health Data Research UK, which is funded by the UK Medical Research Council, Engineering and Physical Sciences Research Council, Economic and Social Research Council, Department of Health and Social Care (United Kingdom), Chief Scientist Office of the Scottish Government Health and Social Care Directorates, Health and Social Care Research and Development Division (Welsh Government), Public Health Agency (Northern Ireland), British Heart Foundation and Wellcome Trust. The Authors acknowledge the ERN Euro-NMD for support. PVD holds a senior clinical investigatorship of FWO-Vlaanderen (G077121N) and is supported by the E. von Behring Chair for Neuromuscular and Neurodegenerative Disorders, the ALS Liga België and the KU Leuven funds “Een Hart voor ALS”, “Laeversfonds voor ALS Onderzoek” and the “Valéry Perrier Race against ALS Fund”. Several authors of this publication are member of the European Reference Network for Rare Neuromuscular Diseases (ERN-NMD).

