## Supplementary materials for "Unsupervised machine-learning identifies clinically distinct subtypes of ALS that reflect different genetic architectures and biological mechanisms"

### 1 Supplementary methods

#### 1.1 Generation of polygenic risk scores

GWAS summary statistics were first processed through a standard data cleaning protocol. We retained only single nucleotide polymorphisms (SNPs), excluding any non-SNP or strand-ambiguous variants. Variants were filtered to those present within the 1000 Genomes phase 3 (1KG) European ancestry population reference (1) (n = 503) and HapMap3 reference panel (2). For the ALS GWAS, where chromosomal positions were not available, SNPs were matched to the 1KG reference panel by rsID. For all other GWAS, SNPs were matched by GRCh37 chromosomal position using *bigsnpr* (version 1.11.6) (3). After matching, allele order was harmonised with the reference.

If not already reported, and where possible, we calculated per-SNP effective sample size (N_eff_) from per-SNP case and control sample sizes (4). If this could not be determined per-SNP, all SNPs were assigned a single N_eff_, calculated as a sum of N_eff_ values for each cohort contributing to the GWAS meta-analysis.

Further processing was performed where possible, excluding SNPs with imputation INFO <0.9, p-values ≤0 or >1, N_eff_ >3 standard deviations from the median N_eff_. We further restricted SNPs to those with minor allele frequency (MAF) >0.01 in both the GWAS and the reference datasets, and excluded any with absolute MAF difference of >0.2 between the two.

PRS were calculated with SBayesR (5) under the reference-standardized approach of *GenoPredPipe* (6); scores for the Project MinE target sample were standardised against those of the 1KG sample. We applied SBayesR using the default settings and the robust parameterisation option. Linkage disequilibrium was estimated using the pre-computed sparse matrices provided, based on 50,000 individuals of European descent from the UK Biobank (5). Where per-SNP N_eff_ was unavailable, we used the impute-n option of SBayesR to exclude variants with imputed N_eff_ >3 standard deviations from the median.

#### 1.2 Clustering of ALS clinical data

Latent class cluster analysis was performed with *Mplus* (version 8.7) and the *MplusAutomation* R package (7) (package version 1.1.0; R version 4.1.3) using the MLR estimator. We attempted to fit latent class models which considered between 1 and 9 latent classes for both the Project MinE discovery sample and the joint sample (pooling together discovery and validation cohorts).

Fitted models were compared first using Akaike (AIC) and Bayesian (BIC) information criterion, where smaller values indicate an improved model fit (8). We additionally considered the theoretical interpretability and parsimony of the accepted solution, which is typical for LCA to minimise identification of small and uninterpretable classes. The quality of the accepted fit was evaluated using entropy and minimum average probability of belonging to the assigned class. High entropy indicates that the identified classes describe the data well (9); values of around 0.8 or higher indicate good model fit. Likewise, greater than 0.8 average probability belonging to the assigned class indicates that people conform well to class assignments.

To ensure that accepted models from each dataset were unbiased by use of full information maximum likelihood to approximate missing values in important model features, the final models were refitted using a subset of the samples, omitting those missing diagnostic delay and disease duration feature information (see Table S4).

In validation of the best-fitting model for the discovery-sample, people from STRENGTH were assigned to classes by *Mplus*.

As a further test of external validity for the accepted model from the discovery sample, we predicted class assignments in STRENGTH using a k-nearest neighbours (KNN) algorithm. The ground truth for class membership was those assigned by *Mplus*. KNN was trained upon the LCA model features using the Project MinE discovery sample (including people with censored disease duration). We allowed KNN to consider between 1 and 20 neighbours, running the algorithm for each value 20 times, and then performing a final prediction for the value with the highest mean accuracy for predicting class assignments in STRENGTH (see Figure S3; Table S5). Area under the receiver operating characteristic curve (AUC) was used to determine predictive performance for each class vs any other class. KNN was implemented using the R *class* package (version 7.3.20)(10), and AUC was analysed using *pROC* (version 1.18.0) (11).

#### 1.3 Clinical characterisation of clusters

Linear discriminant analysis was implemented to predict Class in R using the *MASS* (version 7.3.57) package (10). In the analysis, clinical diagnosis (ALS, PLS, or PMA) was dummy coded with ALS as the reference category. Age of onset and disease duration were standardised to have a mean of 0 and standard deviation of 1. Diagnostic delay was, as before, standardised by country of origin. Sex (male or female) and site of onset (bulbar or other) were not recoded. Relationships between linear discriminant axes and clinical variables were examined using pooled within-group correlations, implemented within the *psych* package (version 2.2.9) (12) *statsBy* function.

Multinomial logistic regression analysis was implemented, via the *nnet* (version 7.3.17) (10) package, using the same predictors and variable coding as in linear discriminant analysis. The *MASS* package *stepAIC* function was applied with using forward and backward feature selection to remove any unimportant features on the basis of AIC.

Linear discriminant and multinomial logistic regression analysis methodologies do not account for censoring in data. To ensure that our application of these techniques was not biased by inclusion of people with censored disease duration, the analyses were performed both including and excluding these individuals.

Time-to-event/survival analysis was performed with *survival* (version 3.3.1) (13) *and survminer* (version 0.4.9) (14). Class was first used as a univariate predictor of disease duration and differences were compared using pairwise log-rank tests. A Cox proportional-hazards model was fitted using class and the other clinical variables to predict disease duration. Continuous predictor variables were standardised as before. Disease duration was not standardised in the survival analyses.

#### 1.4 Biological trends across clusters

##### 1.4.1 Rare variant and PRS analyses

The R *stats* (version 4.1.3) *(15)* package was used to perform Fisher’s exact tests for the rare variant analysis and to fit binary logistic regression models for the PRS analyses*.* Odds ratios for class vs other comparisons in the rare variant analyses were derived using the *epitools* (version 0.5.10.1) (16) package *oddsratio.fisher* function.

##### 1.4.2 Gene expression analysis

The processing pipeline used to generate the raw counts expression matrix is available at <https://github.com/rkabiljo/RNASeq_Genes_ERVs>. Briefly, paired FASTQ files were interleaved using BBMap reformat v38.18.0 under default options before adapters were right-clipped and both sides of each read were quality-trimmed with BBMap bbduk v38.18.0. Interleaved files were aligned to hg38 using STAR v2.7.10a (17) before transcripts were quantified using HTSeq (18). Differential expression between assigned classes was performed using DESeq2 (version 4.1.1) (19), controlling for sex, age at death, post-mortem delay, RNA integrity number, and surrogate variables. The scripts used to generate this is available at <https://github.com/rkabiljo/DifferentialExpression_Genes>. Multiple testing correction of differential expression results was performed using independent hypothesis weighting, with an adjusted p-value of <0.05 denoting significance.

##### 1.4.3 Gene enrichment analysis

Gene enrichment analysis was performed using *gprofiler2* (version 4.1.3) (20) and the following databases: Gene Ontology (Biological Process (GO:BP), Molecular Function (GO:MF) and Cellular Component (GO:CC)), Kyoto Encyclopedia of Genes and Genomes (KEGG), Reactome, CORUM, TRANSFAC and miRTarBase. The default g:SCS algorithm was used to assess significant enrichments, with the brain expressed gene expression matrix (~34,000 genes) used as a custom gene background.

#### 1.5 Prediction of cluster membership using baseline data

Random forest and eXtreme Gradient Boosting classification algorithms were trained to predict class membership. Feature importance was evaluated across each trained algorithm based on SHapley Additive exPlanations (SHAP) (24, 25). In a multi-class algorithm, SHAP values can be determined for each level of the multiclass outcome variable and indicate feature importance for predicting a certain group relative to all other groups. A binary classification objective must be used to evaluate feature importance for predictions distinguishing between two specific groups.

Accordingly, a total of 12 machine-learning algorithms were trained. Six algorithms were trained with a multiclass objective, across all classes with sufficient data available in the 3 defined data configurations and two machine-learning approaches applied. A further 6 were trained with a binary classification objective, restricting to people in Classes 1 and 2 only for the 3 data configurations and two machine-learning approaches.

We evaluated the performance of the multiclass objective algorithms only as the primary objective was the classification across all clusters. Algorithms trained with both the multiclass and binary objectives were evaluated for the secondary investigation of feature importance.

##### 1.5.1 Machine-learning algorithm training procedure

Only people with no missingness on included features were used when training the algorithms. Therefore, the total sample size under the multiclass (binary) objective was 12,508 (11,109) for data configuration 1, and 3,226 (2,990) for data configurations 2 and 3. For data configurations 2 and 3, Class 5 was excluded from the multiclass objectives as fewer than 20 people in the class remained in the sample.

The clinical diagnosis variable was entered into algorithms using one-hot encoding to represent each level across 3 binary features. Diagnostic delay was, as for other analyses, standardised per-country. No other variables were recoded.

Classification algorithms were trained using *caret* (v6.0.93) (26), *randomForest* (v4.7.1.1) (27), and *xgboost* (v1.7.1.1) (28) R packages. Training was primarily performed within the caret *train* function, to maximise the area under the receiver operating characteristic curve (AUC), as calculated within the *multiClassSummary* function for the multiclass objective and within the *twoClassSummary* function for the binary objective (where the metric is labelled ‘ROC’). Class weights in each set of training data were passed to the algorithm to account for class imbalance. Algorithms were trained with 10-fold cross-validation, repeated 10 times.

Repeated cross-validation folds were generated via the *caret* *createMultiFolds* function which uses stratified subsampling across the groups. The cross-validation resamples were regenerated for each stage of parameter tuning with a different fixed seed to ensure pseudo-randomisation replicability.

*Random forest tuning*

Random forests were tuned via grid-search (see Table S10 for the best hyperparameter configuration), using the hyperparameters *ntrees*, *nodesize,* and *mtry* (29).

The *ntrees* hyperparameter was tested at two values, 501 and 1001. This was done to ensure that setting 501 trees was sufficient for classification such that further increases to the number of trees did not substantially improve performance. *nodesize* indicates the minimum number of observations in the terminal node and was tested at all integers between 1 (the default) and 40. The *mtry* values tested varied by model, ranging between $mtry=1$ and $mtry=N_{features}$. Therefore, in the clinical data only forests, mtry was tested at every integer between 1 and 7, and in the forests combining clinical and genetic features, every integer between 1 and 17.

*eXtreme Gradient Boosting tuning*

eXtreme Gradient Boosting algorithms were tuned with the *xgbTree* classifier via a multi-step grid-search (see Table S10 for the best hyperparameter configuration), tuning the hyperparameters *max_depth*, *min_child_weight*, *gamma*, *subsample*, *colsample_bytree*, *eta*, and *nrounds*. Multiclass objectives were trained with the *multi:softprob setting,* and *binary:logistic* was used for binary objectives.

In eXtreme Gradient Boosting, it is important to control the number of boosting iterations, the *nrounds* parameter, performed by the model at a given learning rate, *eta*, to avoid overfitting to the training sample. Therefore, before the first grid search step, we determined an appropriate *nrounds* for the given data at a reasonably high learning rate (*eta* = 0.3). This was performed using the *xgboost* package *xgb.cv* function and the option for early stopping after further iterations no longer improve model performance in the out-of-fold test data. We set this to stop after 50 rounds with no improvement in AUC as calculated by *xgb.cv* across the 10-fold cross validation dataset and compared the optimum *nrounds* across the 10 resamples, using the 3^rd^ Quartile *nrounds,* rounded up to the nearest integer, across resamples for the subsequent grid search steps: $nrounds_{init}$. The other parameters were set to their defaults: *max_depth* = 6, *min_child_weight* = 1, *gamma* = 0.0, *subsample* = 1, *colsample_bytree* = 1.

We next tuned the tree-based parameters (*max_depth*, *min_child_weight*, and *gamma*) via grid search within the *caret* package *train* functionality, holding *subsample* and *colsample_bytree* at their defaults and using *eta* = 0.3 and $nrounds=nrounds_{init}$. Initial grid search was performed across the values: *max_depth* = [5, 6, 7, 8, 9, 10], *min_child_weight* = [1, 2, 3, 4, 5, 6], *gamma* = [0.0, 0.1, 0.2, 0.3, 0.4]. If an optimum parameter value was identified at the edge of the grid search (and not at the limit for that parameter) a smaller subsequent search was performed, recursively extending any applicable parameters by several additional steps outside the values tested until the optimum was found.

The parameters *subsample* and *colsample_bytree* were next trained using the *train* function, taking the initial values of [0.6, 0.65, 0.70, …, 1] for each, setting *eta* = 0.3 and $nrounds=nrounds_{init}$, and lastly *max_depth*, *min_child_weight*, and *gamma* to the optimum values identified prior. As before, the grid search was recursively adjusted when optimum parameters were found at the edge of the grid search.

Finally, the parameters *eta* and *nrounds* were tuned together in a 2-step process, holding all other parameters at the optimum determined prior. First, using *xgb.cv* and the early stopping functionality as before, we determined the optimum number of *nrounds* for each of *eta* = [0.01, 0.02, 0.04, 0.06, 0.08, …, 0.3] as the median *nrounds* across the 10 cross-validation resamples rounded up to the nearest integer. Second, for each pairing of *nrounds* and *eta* we assessed in the *train* function which provided the optimum performance; recursive grid search adjustment was not performed here. The best tuning parameters at this stage were accepted as the optimum tuning for eXtreme Gradient Boosting.

##### 1.5.2 Assessment of model performance and feature importance

Metrics of sensitivity, specificity, precision, and balanced accuracy were calculated via the *caret* package *confusionMatrix* function*.* Receiver operator characteristic curves were generated and the area under the curve calculated using *pROC (11).* In multiclass objectives, curves were generated for each group vs all other groups and for all pairwise group combinations. In binary objectives, a single curve was generated using Class 1 as the reference group and Class 2 as the ‘positive’ value; the other performance metrics were also encoded in this direction.

Feature importance was determined using SHAP values calculated via the R package *kernelshap* (v0.3.3) (30). In multiclass objective algorithms, absolute mean SHAP values were estimated for prediction of each outcome category based on classification probabilities across the full training sample. In binary objectives, absolute mean SHAP values were estimated for classification probability across the overall algorithm. A pseudorandomised subset of approximately 500 records were extracted from the dataset using stratified sampling via the *caret* *createDataPartition* function and supplied as background data required by *kernelshap*; class imbalance was accounted for in *kernelshap* by weighting these data according to class. SHAP values were visualised using *ggplot2*.

### 2 Supplemental figures

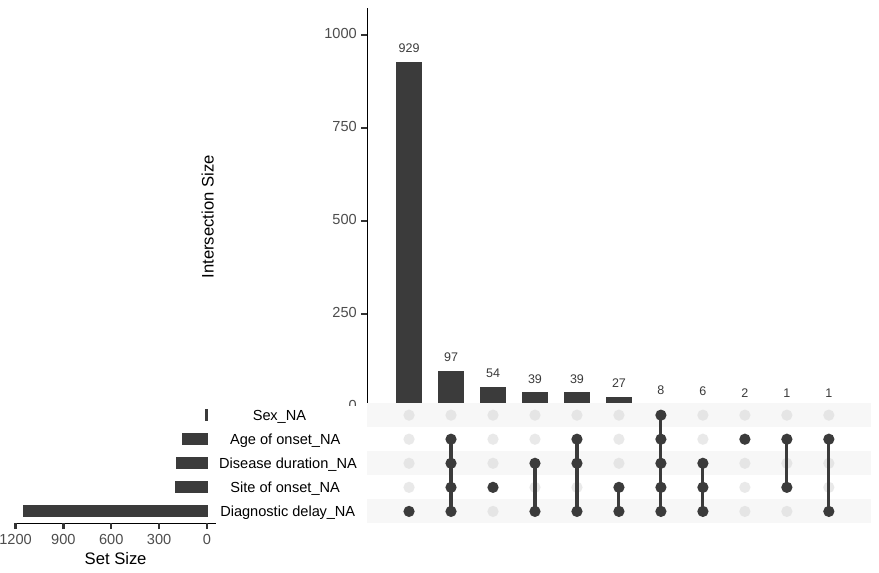

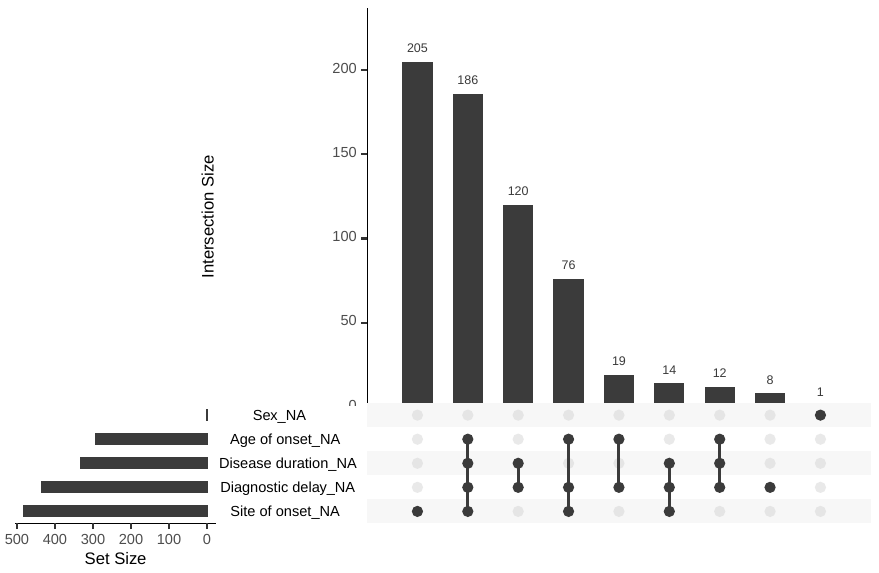

Figure S1. Upset plots for missingness across clinical features used in LCA for the Project MinE (top) and STRENGTH (bottom) cohorts

Plotting was performed with the R naniar package (version 0.6.1)(31).

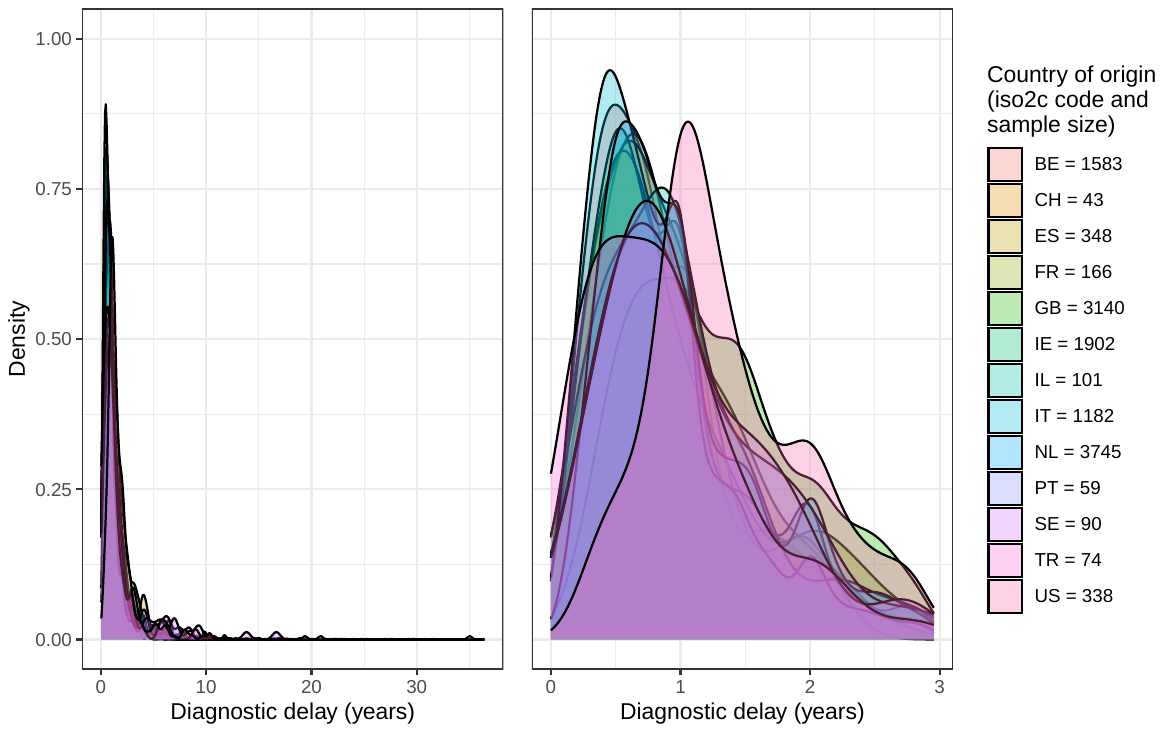

Figure S2. Density plot for diagnostic delay across countries for the combined Project MinE and STRENGTH datasets. The two panels display the same data; the left panel visualises the full distribution of diagnostic delay, and the right panel x-axis is truncated to exclude the final decile of records.

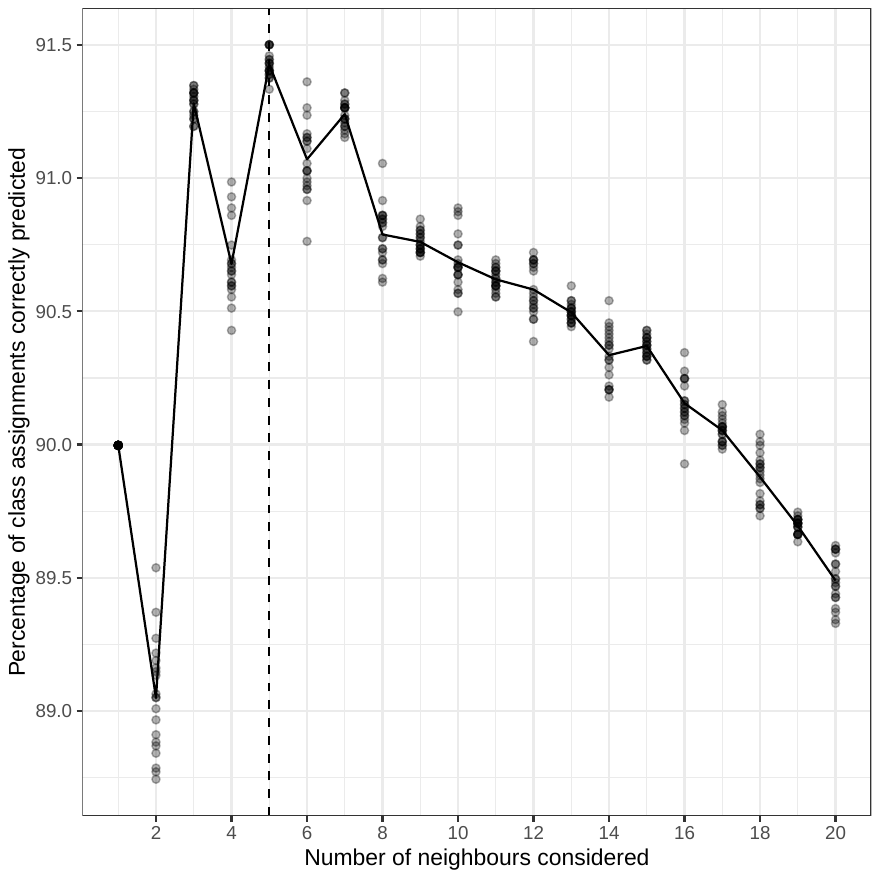

Figure S3. Accuracy of K-Nearest Neighbours algorithm for predicting class membership in STRENGTH when considering between 1 and 20 neighbours

Points indicate performance for each of 20 runs of the KNN algorithm for each number of neighbours using pseudorandomised seeds. The line indicates mean performance for each number of neighbours. The hatched vertical line indicates the configuration with the highest mean predictive accuracy.

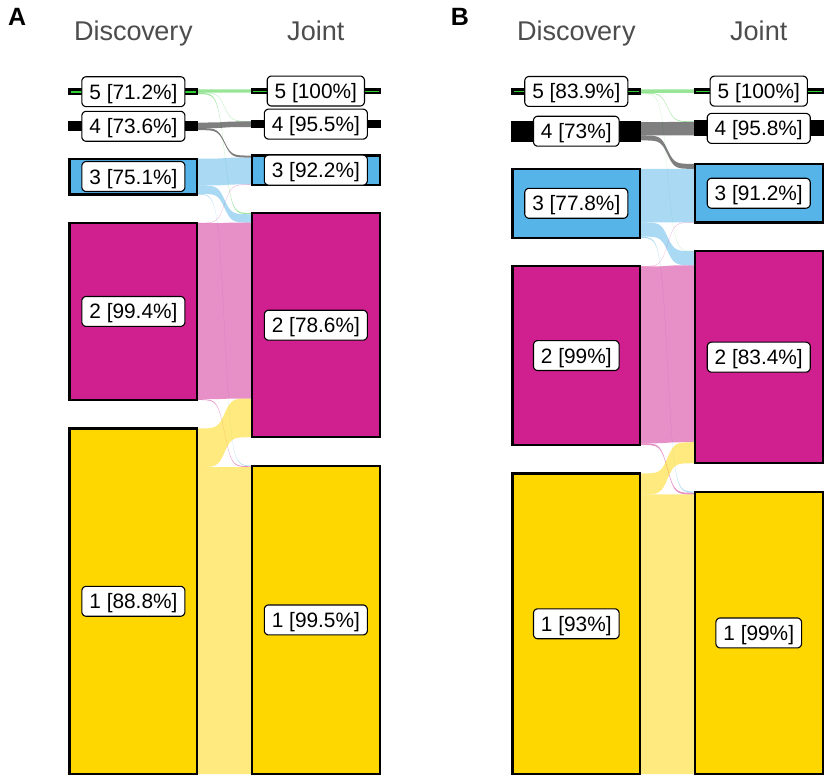

Figure S4. Distribution of people across clusters of the 5-class models fitted to the discovery (Project MinE) and joint (Project MinE and STRENGTH) datasets

Project MinE dataset is shown in Panel A and STRENGTH is shown in Panel B. ‘flows’ on the figure indicate the movement of people between classes across the two models. Percentages shown indicate the proportion of people in a given class who remained in the equivalent class for the model fitted to the opposing dataset (e.g., for Panel A, 88.8% of people from Class 1 of the discovery dataset model remained in Class 1 of the joint dataset model). Plotting was performed using ggsankey (version 0.0.99999)(32).

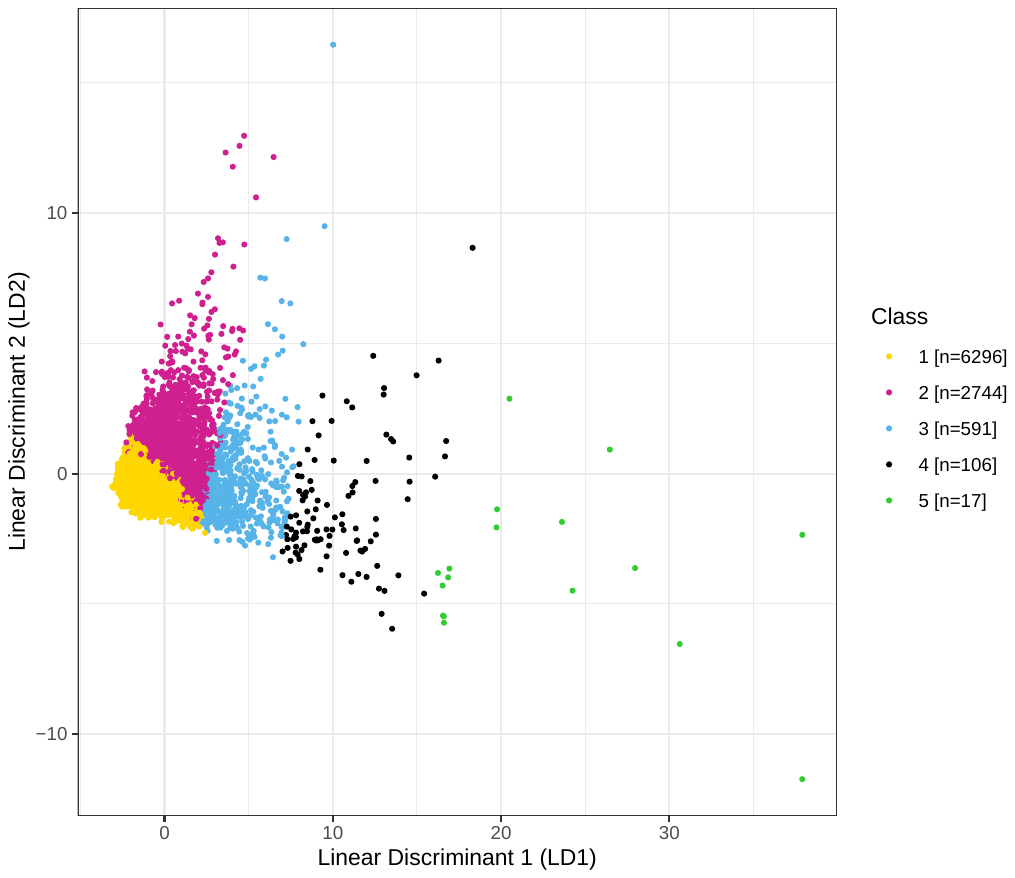

Figure S5. Distribution of people across the first two discriminant axes of linear discriminant analysis for case-complete data restricting to people with non-censored disease duration in the joint dataset

LD1 is highly correlated with diagnostic delay, while LD2 is associated primarily with disease duration (see

Table S6).

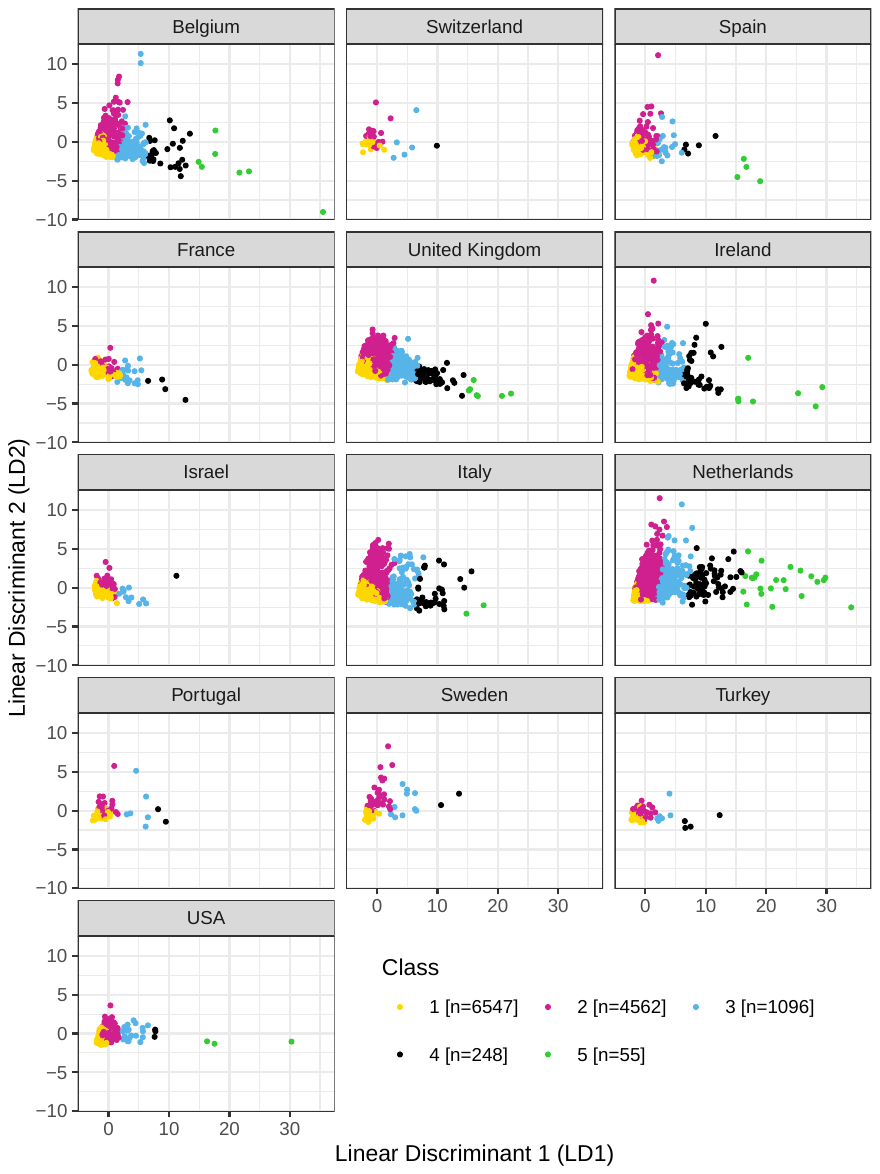

Figure S6. Distribution of people across the first two discriminant analysis axes for people with no missing data in the joint dataset, stratified by country of origin

LD1 is highly correlated with diagnostic delay while LD2 is associated primarily with disease duration (see Table 3).

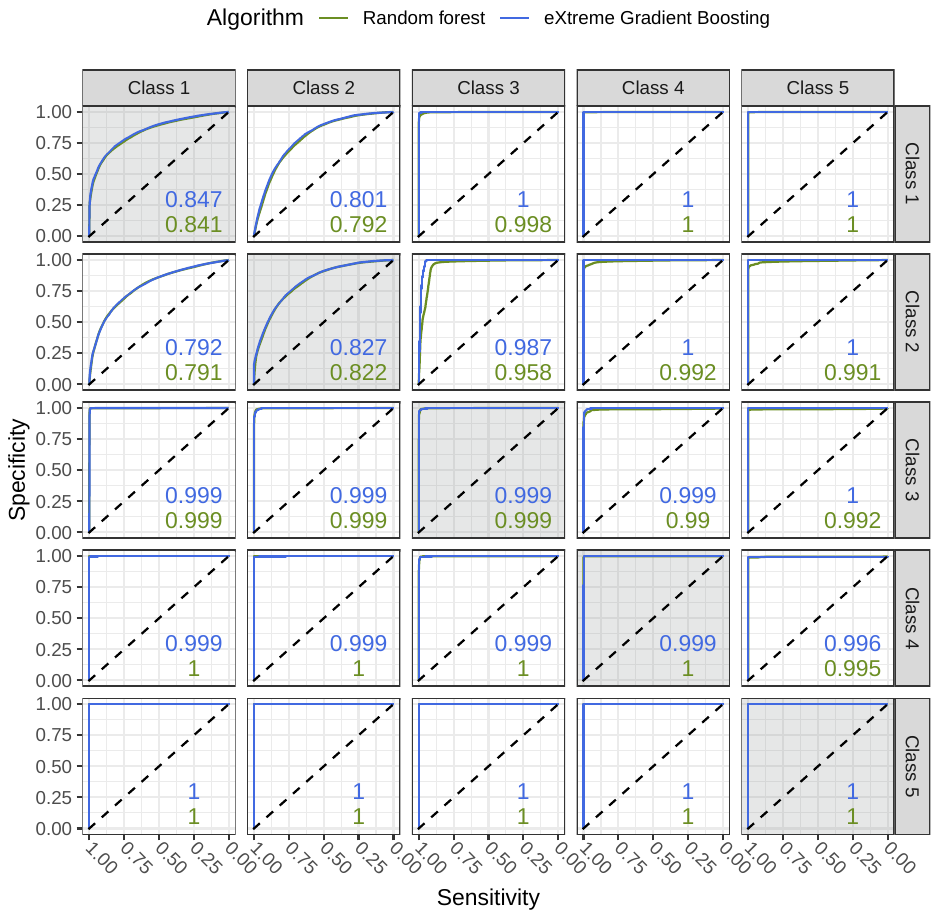

Figure S7. Receiver operating characteristic curves for random forest and extreme gradient boosting algorithm predictions of class membership using only clinical data available around the time of diagnosis across all people with complete clinical data

Panels along the plot-diagonal (displayed with background shading) display ROCs for people being in that class versus all other classes. The upper triangle of panels present ROCs where the class represented in the panel row are considered controls and the column for the class is the ‘case’ group. Case-control coding is reversed for panels in the lower triangle. Colour denotes performance of different machine-learning algorithms, with numbers shown representing the area under the curve for each algorithm in that panel.

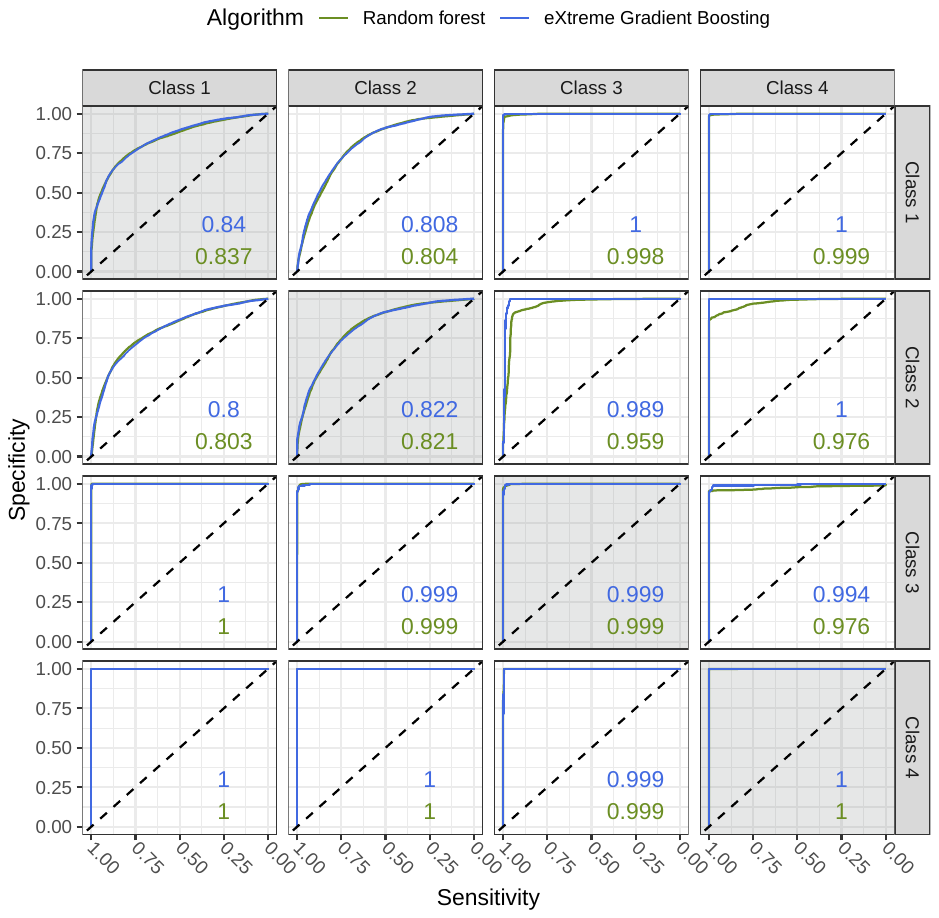

Figure S8. Receiver operating characteristic curves for random forest and extreme gradient boosting algorithm predictions of class membership using clinical data available around the time of diagnosis and measures of genetic disease liability

Panels along the plot-diagonal (displayed with background shading) display ROCs for people being in that class versus all other classes. The upper triangle of panels present ROCs where the class represented in the panel row are considered controls and the column for the class is the ‘case’ group. Case-control coding is reversed for panels in the lower triangle. Colour denotes performance of different machine-learning algorithms, with numbers shown representing the area under the curve for each algorithm in that panel. Class 5 is excluded from this classification analysis owing to small sample size.

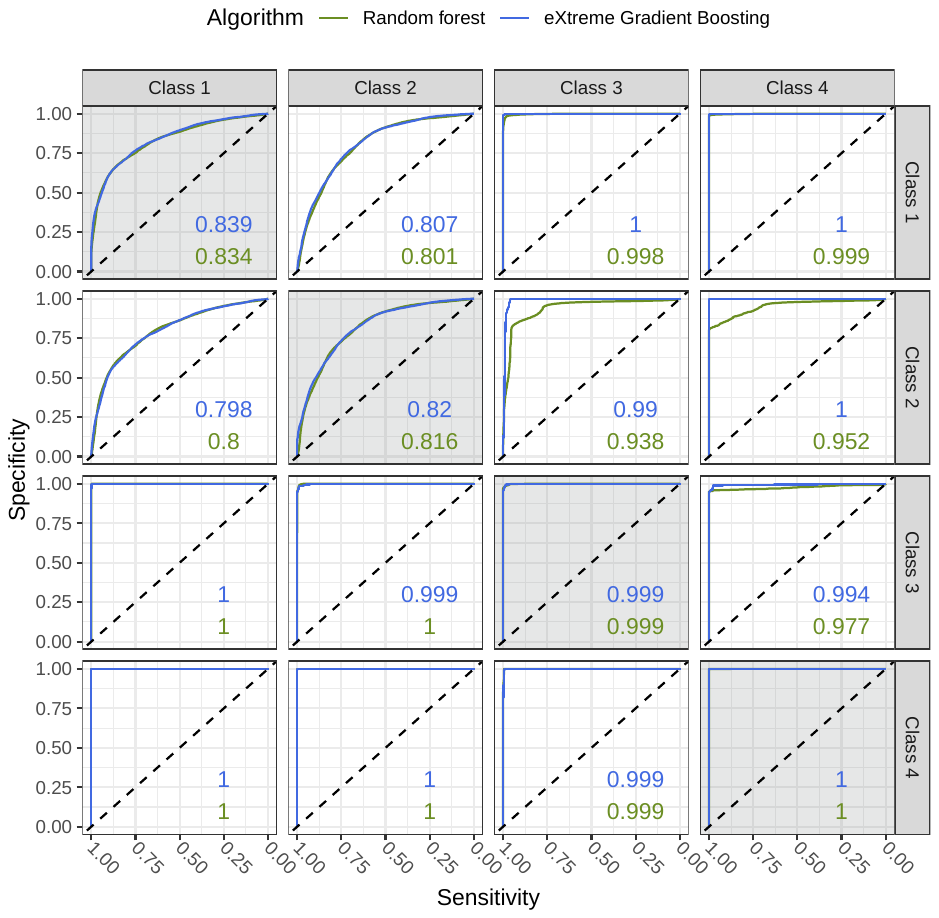

Figure S9. Receiver operating characteristic curves random forest and extreme gradient boosting algorithm predictions of class membership using clinical data available around the time of diagnosis

This classification algorithm was trained upon the same features as the algorithm presented in Figure S7, but was restricted to the same samples that were used in the algorithm presented in Figure S8. Panels along the plot-diagonal (displayed with background shading) display ROCs for people being in that class versus all other classes. The upper triangle of panels present ROCs where the class represented in the panel row are considered controls and the column for the class is the ‘case’ group. Case-control coding is reversed for panels in the lower triangle. Colour denotes performance of different machine-learning algorithms, with numbers shown representing the area under the curve for each algorithm in that panel. Class 5 is excluded from this classification analysis owing to small sample size.

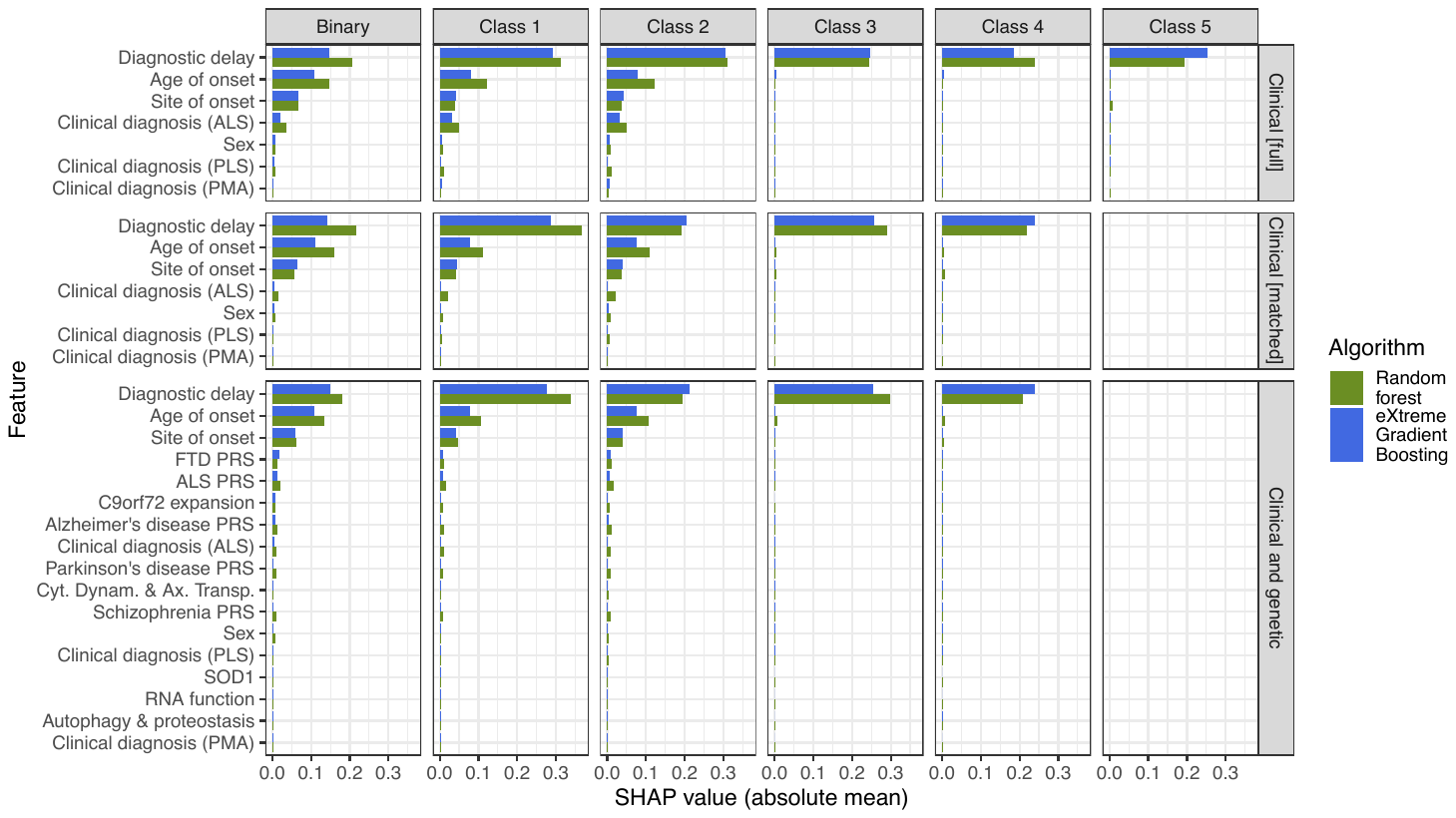

Figure S10. SHapley Additive exPlanations (SHAP) of feature importance across trained classification algorithms

Panel rows stratify between algorithms using ‘Clinical’ features only, and with a combination of ‘Clinical and genetic’ features; the ‘Clinical [full]’ row is trained upon all samples with complete clinical data, while ‘Clinical [matched]’ uses the same features but is sample matched to the ‘Clinical and genetic’ row. Panel columns stratify feature importance for predictions of probability of each class individually; the ‘Binary’ column describes distinct algorithms trained to predict classes 1 versus 2 only. Class 5 is only represented within Clinical [all] algorithms. FTD = frontotemporal dementia, ALS = amyotrophic lateral sclerosis, PRS = polygenic risk score. SHAP values are calculated for predicted class probabilities and therefore can range between 0 and 1. Figures SX1-S12 give greater detail about the distribution SHAP values across data.

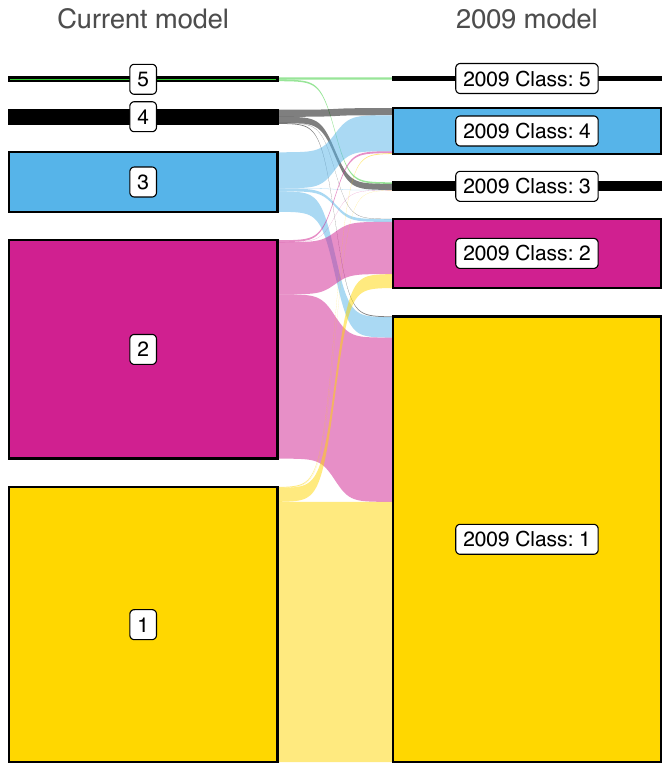

Figure S11. Comparison of class assignments for people from STRENGTH (N = 5,961) within the (joint dataset) 5-class model and a 5-class model identified through latent class analysis within a previous study(33)

The left column indicates class assignments by current model and the right column is those from the previous study (‘2009 model’). Colouring of the 2009 model column is with respect to the current model class with the largest sample overlap.

### 3 Supplemental tables

Table S1. Mean and standard deviation of diagnostic delay per country of origin across unique samples from Project MinE and STRENGTH

Diagnostic delay values entered into the latent class model and subsequent analyses were centred on the per-country mean and scaled by the per-country standard deviation. The values presented for ‘total’ diagnostic delay were derived by aggregating across the per-country statistics weighted by sample size; these were not used in any analyses.

| Region (Iso2c code) | Number of samples with measured diagnostic delay | Mean diagnostic delay in years | Standard deviation |
| --- | --- | --- | --- |
| Belgium (BE) | 1583 | 1.111 | 1.257 |
| Switzerland (CH) | 43 | 1.357 | 1.298 |
| Spain (ES) | 348 | 1.142 | 1.334 |
| France (FR) | 166 | 1.123 | 0.967 |
| United Kingdom (GB) | 3140 | 1.711 | 1.461 |
| Ireland (IE) | 1902 | 1.238 | 1.347 |
| Israel (IL) | 101 | 1.191 | 1.153 |
| Italy (IT) | 1182 | 0.935 | 0.779 |
| Netherlands (NL) | 3745 | 1.669 | 2.609 |
| Portugal (PT) | 59 | 1.695 | 1.830 |
| Sweden (SE) | 90 | 2.187 | 2.872 |
| Turkey (TR) | 74 | 1.387 | 1.584 |
| United States of America (US) | 338 | 2.035 | 2.782 |
| **Total** | **12771** | **1.464** | **1.872** |

Table S2. Summary of number of variants identified in ALS-associated genes and assignment of genes to disease pathways according to evidence of role in gene products in pathway

Additional pathways relevant to ALS but without any variants assigned are: DNA repair (D), mitochondrial dysfunction (E), inflammation (F)

| Assigned to Pathway | Gene | Number of people with variants in gene | Supporting evidence for role in pathway | Other pathway involvement |
| --- | --- | --- | --- | --- |
| A – Autophagy and proteostasis | *UBQLN2* | *11* | *(34-36)* | *B (34)* |
|  | *CHMP2B* | *23* | *(34, 37)* | *A (38), C (38-40)* |
|  | *ERBB4* | *62* | *(34, 41)* | *E (39)* |
|  | *SIGMAR1* | *17* | *(34, 42)* | *E (42)* |
|  | *CHCHD10* | *22* | *(40, 43)* | *E (34, 40, 43)* |
|  | *DAO* | *49* | *(44)* |  |
|  | *OPTN* | *67* | *(34, 36, 37, 43, 45)* | *F (40)* |
|  | *SCFD1* | *47* | *(46)* |  |
|  | *SQSTM1 (p62)* | *51* | *(34, 36, 37)* | *F (40)* |
|  | *TBK1* | *96* | *(34, 36, 43, 47)* | *F (34, 39, 40, 43)* |
|  | *UNC13A* | *88* | *(48)* | *C (49)* |
|  | *VAPB* | *12* | *(34, 50)* | *C (34, 39, 51), D (43)* |
|  | *VCP* | *22* | *(34, 36-38, 52)* |  |
|  | *VEGFA* | *41* | *(45, 53)* |  |
| B – RNA function | *ANG* | *26* | *(34, 54)* | *A (34, 39, 40, 45)* |
|  | *ATXN2* | *177* | *(36, 55)* | *C (39)* |
|  | *FUS* | *52* | *(34, 36)* | *A (38), D (37, 43), E (34)* |
|  | *hnRNPA1* | *10* | *(36, 43, 56)* |  |
|  | *MATR3* | *46* | *(34, 36, 40)* | *A (57)* |
|  | *SETX* | *213* | *(34, 58)* |  |
|  | *TAF15* | *62* | *(34)* |  |
|  | *TARDBP* | *23* | *(34, 59)* | *A (38), D (43)* |
| C – Cytoskeletal dynamics and axonal transport | *ALS2* | *104* | *(37, 60)* | *D (43)* |
|  | *ANXA11* | *67* | *(43, 61)* |  |
|  | *CFAP410 (C21ORF2)* | *78* | *(40, 43)* | *A (57), D (37, 40, 62), E (40)* |
|  | *FIG4* | *80* | *(34, 38)* |  |
|  | *NEFH* | *114* | *(37, 60, 63)* |  |
|  | *NEK1* | *160* | *(40, 43)* | *A (57), D* (37, 40, 62) *, E (40, 43)* |
|  | *ATXN1* | *64* | *(64, 65)* |  |
|  | *DCTN1* | *100* | *(37, 60)* |  |
|  | *MOBP* | *6* | *(66-68)* |  |
|  | *PFN1* | *6* | *(37, 60)* | *A (60), B (60)* |
|  | *SPG11* | *236* | *(34, 38, 69)* | *A (38), D (37)* |
|  | *TUBA4A* | *7* | *(37, 60)* |  |
| Not assigned to pathway | *SOD1* | *53* | *(70)* | *-* |
|  | *C9orf72* | *366* | *(71)* | *-* |

Table S3. Comparison of latent class model solutions for the Project MinE and for the Joint datasets

The joint dataset is a combination of unique individuals from Project MinE and STRENGTH. AIC = Akaike information criterion, (a)BIC = (adjusted) Bayesian information criterion

| Dataset |  | 1 | 2 | 3 | 4 | 5 | 6 | 7 | 8 | 9 |
| --- | --- | --- | --- | --- | --- | --- | --- | --- | --- | --- |
| Project MinE (discovery sample) | Loglikelihood | -50036.833 | -48440.246 | -47557.668 | -46908.637 | -46502.638 | - | - | - | - |
|  | AIC | 100089.667 | 96912.492 | 95163.337 | 93881.274 | 93085.276 | - | - | - | - |
|  | BIC | 100143.932 | 97021.022 | 95326.131 | 94098.333 | 93356.6 | - | - | - | - |
|  | aBIC | 100118.51 | 96970.178 | 95249.865 | 93996.645 | 93229.49 | - | - | - | - |
|  | Entropy | 1 | 0.937 | 0.912 | 0.787 | 0.791 | - | - | - | - |
|  | N per class | 6523 | 243: 6280 | 415: 109: 5999 | 318: 81: 2096: 4028 | 3952: 2023: 409: 87: 52 | - | - | - | - |
|  | Lowest average class probability | NA | 0.877 | 0.802 | 0.843 | 0.818 | - | - | - | - |
| Joint | Loglikelihood | -100154.4 | -96218.178 | -94005.047 | -92337.947 | -91200.445 | -90418.286 | -90000.826 | -89507.894 | -89228.181 |
|  | AIC | 200324.807 | 192468.356 | 188058.095 | 184739.894 | 182480.889 | 180932.572 | 180113.652 | 179143.788 | 178600.362 |
|  | BIC | 200385.38 | 192589.502 | 188239.814 | 184982.187 | 182783.755 | 181296.011 | 180537.664 | 179628.373 | 179145.52 |
|  | aBIC | 200359.957 | 192538.655 | 188163.544 | 184880.493 | 182656.638 | 181143.472 | 180359.701 | 179424.987 | 178916.71 |
|  | Entropy | 1 | 0.954 | 0.933 | 0.82 | 0.836 | 0.841 | 0.79 | 0.792 | 0.752 |
|  | N per class | 14352 | 524: 13828 | 136: 1100: 13116 | 5400: 108: 717: 8127 | 7401: 5470: 1138: 259: 84 | 1260: 329: 47: 88: 5346: 7282 | 1395: 326: 6601: 1263: 23: 92: 4652 | 404: 1408: 55: 16: 1471: 139: 6172: 4687 | 1254: 4269: 3032: 1387: 55: 3798: 138: 404: 15 |
|  | Lowest average class probability | NA | 0.926 | 0.894 | 0.884 | 0.879 | 0.828 | 0.748 | 0.738 | 0.711 |

Table S4. Five-class latent class model solutions when restricting to samples with recorded diagnostic delay and disease duration

Numbers presented in bold refer to average posterior probability of belonging to the assigned class. ^†^Total percentage of people in the equivalent classes across the missingness vs full models were: 99.2% for the discovery model; 99.6% for the joint dataset model.

| Dataset  [General statistics] | Assigned class | Percentage of class in equivalent class of full-dataset model ^†^ | N in class (% of dataset) based on | | Average posterior probability of belonging to class | | | | |
| --- | --- | --- | --- | --- | --- | --- | --- | --- | --- |
|  |  |  | **posterior probabilities** | **most likely class membership** | **1** | **2** | **3** | **4** | **5** |
| Discovery  (Project MinE)  [N = 5377; entropy = 0.850] | 1 | 99.0 | 3156.30 (0.587) | 3319 (0.617) | **0.908** | 0.09 | 0.002 | 0 | 0 |
|  | 2 | 99.7 | 1699.04 (0.316) | 1551 (0.288) | 0.089 | **0.883** | 0.028 | 0 | 0 |
|  | 3 | 99.3 | 422.66 (0.079) | 408 (0.076) | 0.008 | 0.075 | **0.909** | 0.007 | 0 |
|  | 4 | 97.6 | 82.99 (0.015) | 83 (0.015) | 0 | 0 | 0.037 | **0.963** | 0 |
|  | 5 | 1 | 16.01 (0.003) | 16 (0.003) | 0 | 0 | 0 | 0 | **1** |
| Joint  [N = 12771; entropy = 0.881] | 1 | 99.4 | 6348.46 (0.497) | 6624 (0.519) | **0.923** | 0.076 | 0.001 | 0 | 0 |
|  | 2 | 99.9 | 4949.39 (0.388) | 4698 (0.368) | 0.049 | **0.926** | 0.025 | 0 | 0 |
|  | 3 | 99.9 | 1153.11 (0.090) | 1132 (0.089) | 0.003 | 0.084 | **0.902** | 0.011 | 0 |
|  | 4 | 99.6 | 259.75 (0.020) | 257 (0.020) | 0 | 0 | 0.038 | **0.961** | 0.001 |
|  | 5 | 1 | 60.30 (0.005) | 60 (0.005) | 0 | 0 | 0 | 0 | **1** |

Table S5. Validation of the Project MinE dataset 5-class model solution using independent data from STRENGTH within a K-nearest neighbours (KNN) classification algorithm

Class assignments were determined by MPlus, according to the parameters of the 5-class model fitted to the Project MinE dataset. KNN was trained to predict class membership using the clinical features of LCA. The training dataset for KNN was complete cases from the Project MinE sample (N=5320), and prediction was for complete cases (N=7188) from STRENGTH. Area under the receiver operating characteristic curve (AUC) for the KNN model in prediction of each assigned class vs any other class was consistently high. Numbers presented in bold denote people predictions which align with the assigned class. The algorithm was applied in R using a fixed seed, and 5 neighbours were considered (see Figure S3).

| Class assigned by latent class model | Number of people predicted to belong to class by KNN | | | | | AUC for KNN prediction of assigned class vs other |
| --- | --- | --- | --- | --- | --- | --- |
|  | **1** | **2** | **3** | **4** | **5** |  |
| 1 | **3646** | 110 | 5 | 0 | 0 | 0.938 |
| 2 | 300 | **1907** | 19 | 0 | 0 | 0.906 |
| 3 | 17 | 111 | **770** | 11 | 0 | 0.919 |
| 4 | 0 | 0 | 32 | **209** | 0 | 0.932 |
| 5 | 0 | 0 | 0 | 11 | **40** | 0.892 |

Table S6. Results of linear discriminant analysis after restricting to people with non-censored disease duration (N = 9,754)

Proportion of trace describes the proportion of the separation between classes accounted for by each linear discriminant (LD) axis. Pooled within-group correlations greater than 0.5 are presented in bold and are considered variables associated with a given LD. Reference groups for categorical variables are: ‘not-bulbar’ for site of onset, ‘male’ for sex, ‘ALS’ for clinical diagnosis. Figure S5 visualises the distribution of people and classes across the first two LD axes.

| Statistic | Variable | LD1 | LD2 | LD3 | LD4 |
| --- | --- | --- | --- | --- | --- |
| Eigenvalue | - | 95.48 | 31.93 | 3.80 | 2.16 |
| Proportion of trace | - | 0.898 | 0.100 | 1.42E-03 | 4.59E-04 |
| Pooled within-group correlation | Diagnostic delay | **0.923** | -0.375 | -0.082 | -0.001 |
|  | Age of onset | -0.067 | -0.342 | -0.353 | **-0.580** |
|  | Disease duration | **0.507** | **0.800** | 0.089 | -0.084 |
|  | Site of onset (bulbar) | -0.073 | -0.264 | 0.475 | 0.177 |
|  | Sex (female) | -0.012 | -0.081 | -0.208 | -0.214 |
|  | Clinical diagnosis (PLS) | 0.099 | 0.013 | **0.699** | **-0.646** |
|  | Clinical diagnosis (PMA) | 0.030 | 0.123 | -0.268 | -0.386 |

Table S7. Results of multinomial regression analysis of all people with no missingness across predictors, including people with censored disease duration

Class 1 is used as the outcome variable reference category because this is the largest subgroup. Continuous variables were standardised to have a mean of 0 and standard deviation of 1; diagnostic delay is standardised per-country of origin. Categorical variable reference categories are: ‘not-bulbar’ for site of onset; “ALS” for clinical diagnosis. Sex at birth (male or female) was entered into the regression model but removed in stepwise feature selection. ALS = amyotrophic lateral sclerosis, PLS = primary lateral sclerosis, PMA = progressive muscular atrophy.

| Class | Predictor | Standardised  beta (95% Confidence Interval) | Standard error | Z-score | p-value |
| --- | --- | --- | --- | --- | --- |
| 2 | Diagnostic delay | 1.18 (0.98, 1.4) | 0.104 | 11 | 5.05E-30 |
|  | Age of onset | -0.743 (-0.83, -0.66) | 0.0442 | -17 | 2.83E-63 |
|  | Disease duration | 9.72 (9.3, 10) | 0.225 | 43 | 0 |
|  | Site of onset (bulbar) | -1.04 (-1.2, -0.88) | 0.0849 | -12 | 1.14E-34 |
|  | Clinical diagnosis (PLS) | 2.5 (1.8, 3.2) | 0.344 | 7.3 | 4.04E-13 |
|  | Clinical diagnosis (PMA) | 1.44 (1.1, 1.8) | 0.179 | 8.1 | 7.52E-16 |
| 3 | Diagnostic delay | 41.4 (34, 49) | 3.96 | 10 | 1.50E-25 |
|  | Age of onset | -0.0527 (-0.5, 0.39) | 0.226 | -0.23 | 0.815 |
|  | Disease duration | 11.8 (11, 13) | 0.373 | 32 | 1.05E-221 |
|  | Site of onset (bulbar) | -2.99 (-4, -2) | 0.531 | -5.6 | 1.72E-08 |
|  | Clinical diagnosis (PLS) | 9.23 (7.2, 11) | 1.04 | 8.9 | 6.91E-19 |
|  | Clinical diagnosis (PMA) | 2.86 (1.4, 4.3) | 0.757 | 3.8 | 0.000162 |
| 4 | Diagnostic delay | 87.6 (61, 110) | 13.4 | 6.5 | 6.25E-11 |
|  | Age of onset | -1.64 (-3.1, -0.23) | 0.72 | -2.3 | 0.0227 |
|  | Disease duration | 12.9 (12, 14) | 0.629 | 21 | 1.62E-93 |
|  | Site of onset (bulbar) | -5.92 (-13, 0.69) | 3.37 | -1.8 | 0.0791 |
|  | Clinical diagnosis (PLS) | 12.3 (8.3, 16) | 2.02 | 6.1 | 1.13E-09 |
|  | Clinical diagnosis (PMA) | 4.6 (-0.064, 9.3) | 2.38 | 1.9 | 0.0532 |
| 5 | Diagnostic delay | 111 (94, 130) | 8.33 | 13 | 3.38E-40 |
|  | Age of onset | -2.87 (-6.5, 0.74) | 1.84 | -1.6 | 0.119 |
|  | Disease duration | 12.7 (10, 15) | 1.31 | 9.7 | 2.95E-22 |
|  | Site of onset (bulbar) | -11.4 (-51, 28) | 20.3 | -0.56 | 0.573 |
|  | Clinical diagnosis (PLS) | 9.24 (-2.7, 21) | 6.1 | 1.5 | 0.13 |
|  | Clinical diagnosis (PMA) | 0.733 (-16, 17) | 8.39 | 0.087 | 0.93 |

Table S8. Results of multinomial regression analysis of all people with no missingness across predictors, restricted to people with non-censored disease duration

Class 1 is used as the outcome variable reference category because this is the largest subgroup. Continuous variables were standardised to have a mean of 0 and standard deviation of 1; diagnostic delay is standardised per-country of origin. Categorical variable reference categories are: ‘not-bulbar’ for site of onset; “ALS” for clinical diagnosis, ‘male’ for sex. No features were dropped from the model within stepwise feature selection.* No person with PMA remained in this class for this data subsample. ALS = amyotrophic lateral sclerosis, PLS = primary lateral sclerosis, PMA = progressive muscular atrophy.

| Class | Predictor | Standardised  beta (95% Confidence Interval) | Standard error | Z-score | p-value |
| --- | --- | --- | --- | --- | --- |
| 2 | Diagnostic delay | 15.6 (12, 19) | 1.59 | 9.8 | 8.06E-23 |
|  | Age of onset | -6.8 (-8.2, -5.5) | 0.689 | -9.9 | 5.43E-23 |
|  | Disease duration | 213 (170, 250) | 20.2 | 11 | 3.33E-26 |
|  | Site of onset (bulbar) | -10.1 (-12, -8) | 1.07 | -9.5 | 3.12E-21 |
|  | Sex (female) | -2.09 (-3, -1.2) | 0.464 | -4.5 | 6.48E-06 |
|  | Clinical diagnosis (PLS) | 41.5 (33, 50) | 4.23 | 9.8 | 1.06E-22 |
|  | Clinical diagnosis (PMA) | 15.8 (12, 19) | 1.69 | 9.3 | 1.05E-20 |
| 3 | Diagnostic delay | 158 (120, 190) | 18.6 | 8.5 | 2.14E-17 |
|  | Age of onset | -5.74 (-7.4, -4.1) | 0.85 | -6.8 | 1.45E-11 |
|  | Disease duration | 219 (180, 260) | 20.6 | 11 | 2.06E-26 |
|  | Site of onset (bulbar) | -16.9 (-21, -13) | 2.07 | -8.2 | 3.64E-16 |
|  | Sex (female) | -0.0116 (-2, 1.9) | 0.99 | -0.012 | 0.991 |
|  | Clinical diagnosis (PLS) | 66.6 (50, 84) | 8.69 | 7.7 | 1.83E-14 |
|  | Clinical diagnosis (PMA) | 19.5 (14, 25) | 2.59 | 7.5 | 6.15E-14 |
| 4 | Diagnostic delay | 193 (150, 240) | 22 | 8.8 | 1.82E-18 |
|  | Age of onset | -6.95 (-9.6, -4.3) | 1.36 | -5.1 | 3.43E-07 |
|  | Disease duration | 222 (180, 260) | 20.8 | 11 | 9.74E-27 |
|  | Site of onset (bulbar) | -19.8 (-27, -13) | 3.6 | -5.5 | 4.03E-08 |
|  | Sex (female) | 0.768 (-3.1, 4.6) | 1.95 | 0.39 | 0.694 |
|  | Clinical diagnosis (PLS) | 63.1 (41, 85) | 11.3 | 5.6 | 2.10E-08 |
|  | Clinical diagnosis (PMA) | 26.3 (-21, 74) | 24.1 | 1.1 | 0.276 |
| 5 | Diagnostic delay | 200 (160, 240) | 22.3 | 9 | 2.76E-19 |
|  | Age of onset | -7.23 (-15, 0.93) | 4.16 | -1.7 | 0.0826 |
|  | Disease duration | 222 (180, 260) | 20.8 | 11 | 1.92E-26 |
|  | Site of onset (bulbar) | -22.1 (-36, -8.4) | 7 | -3.2 | 1.56E-03 |
|  | Sex (female) | 0.696 (-11, 12) | 5.79 | 0.12 | 0.904 |
|  | Clinical diagnosis (PLS) | 52.6 (24, 81) | 14.4 | 3.6 | 2.67E-04 |
|  | Clinical diagnosis (PMA)* | -23.02 (-) | - | - | - |

Table S9. Summary of cox proportional-hazards model predicting disease duration from onset until death or censoring using Class and all other clinical features from LCA

Continuous predictors (age of onset and diagnostic delay) were standardised to have a mean of 0 and standard deviation of 1; Diagnostic delay was standardised by country of origin (see Table S1; Figure S2). Hazard ratios greater than 1 indicate association between the variable and shorter disease duration.

| Variable | Factor Level | Number sampled | Hazard Ratio [95% Confidence Interval] | Inverse hazard ratio | Z-score | P-value |
| --- | --- | --- | --- | --- | --- | --- |
| Class | 1 | 6547 | - | - | - | - |
|  | 2 | 4562 | 0.010 [0.009, 0.012] | 99.3 | -68 | <2e-16 |
|  | 3 | 1096 | 0.011 [0.010, 0.014] | 87.8 | -48.8 | <2e-16 |
|  | 4 | 248 | 0.016 [0.011, 0.021] | 64.6 | -26.1 | <2e-16 |
|  | 5 | 55 | 0.045 [0.024, 0.086] | 22 | -9.46 | <2e-16 |
| Site of onset | Other | 8736 | - | - | - | - |
|  | Bulbar | 3772 | 0.940 [0.899, 0.983] | 1.06 | -2.7 | 0.00699 |
| Sex | Male | 7412 | - | - | - | - |
|  | Female | 5096 | 0.925 [0.888, 0.964] | 1.08 | -3.68 | 0.000237 |
| Clinical diagnosis | ALS | 11406 | - | - | - | - |
|  | PLS | 473 | 0.327 [0.274, 0.391] | 3.05 | -12.4 | <2e-16 |
|  | PMA | 629 | 0.824 [0.742, 0.915] | 1.21 | -3.63 | 0.000281 |
| Age of onset in years | - | 12508 | 1.21 [1.18, 1.24] | 0.828 | 16 | <2e-16 |
| Diagnostic delay | - | 12508 | 0.644 [0.610, 0.681] | 1.55 | -15.8 | <2e-16 |

Table S10. Optimum hyperparameter tuning settings and overall AUC for all machine-learning algorithms trained

Predictions were made based on ‘Clinical’ features only or a combination of ‘Clinical and genetic’ features; the ‘Clinical [full]’ columns are trained based on samples with complete clinical data, while ‘Clinical [matched]’ uses the same features but is sample matched to the ‘Clinical and genetic’ column In binary classifications, data were further subsampled to only people assigned to classes 1 and 2. AUC values shown in bold indicate the best performing model for a given dataset configuration. Note that the Clinical [full] model describes a multiclass model predicting 5 classes, while the other multiclass models exclude class 5 owing to small sample size. Therefore, these AUC values cannot directly be compared for the multiclass models between Clinical [Full] and with Clinical and genetic or Clinical [matched] datasets. Refer instead to the comparisons shown across Figure S7-Figure S9. ^†^calculated within the R caret package multiClassSummary function for multiclass and twoClassSummary for binary objectives.

|  | | Algorithm objective | | | | | |
| --- | --- | --- | --- | --- | --- | --- | --- |
|  |  | **Multiclass** | | | **Binary (Classes 1 and 2)** | | |
| Dataset | | Clinical [Full] | Clinical and genetic | Clinical [matched] | Clinical [Full] | Clinical and genetic | Clinical [matched] |
| XGboost tuning parameters | nrounds | 172 | 65 | 62 | 769 | 234 | 354 |
|  | max_depth | 4 | 6 | 6 | 3 | 3 | 2 |
|  | eta | 0.18 | 0.1 | 0.16 | 0.02 | 0.02 | 0.02 |
|  | gamma | 0 | 0.1 | 0.1 | 0 | 0.2 | 0.1 |
|  | colsample_bytree | 1 | 0.7 | 0.7 | 0.7 | 0.8 | 0.65 |
|  | min_child_weight | 1 | 4 | 4 | 6 | 9 | 1 |
|  | subsample | 1 | 0.85 | 0.8 | 0.95 | 0.95 | 0.9 |
| Random forest tuning parameters | ntree | 1001 | 1001 | 1001 | 1001 | 1001 | 1001 |
|  | nodesize | 30 | 37 | 32 | 38 | 39 | 40 |
|  | mtry | 4 | 9 | 4 | 3 | 5 | 3 |
| Overall AUC^†^ for prediction across all included classes | eXtreme Gradient Boosting | **0.935** | **0.916** | **0.915** | **0.803** | **0.813** | **0.812** |
|  | Random forest | 0.932 | 0.915 | 0.913 | 0.794 | 0.807 | 0.805 |

12.	Revelle W. psych: Procedures for Personality and Psychological

Research. R package version 2.2.9. ed. Northwestern University, Evanston, Illinois, USA2022.

13.	Therneau T. A Package for Survival Analysis in R. 3.3.1 ed2022.

14.	Kassambara A, Kosinski M, Biecek P. survminer: Drawing Survival Curves using 'ggplot2'. 0.4.9 ed2021.

15.	R Core Team. R: A language and environment for statistical computing. R Foundation for Statistical Computing, Vienna, Austria; 2021.

16.	Aragon TJ. epitools: Epidemiology Tools. 0.5.10.1 ed2020.

24.	Lundberg SM, Lee S-I, editors. A Unified Approach to Interpreting Model Predictions. Adv Neural Inf Process Syst; 2017.

25.	Covert I, Lee S-I. Improving KernelSHAP: Practical Shapley Value Estimation Using Linear Regression. In: Arindam B, Kenji F, editors. Proceedings of The 24th International Conference on Artificial Intelligence and Statistics: PMLR; 2021. p. 3457--65.

26.	Kuhn M. caret: Classification and Regression Training. 6.0.93 ed2022.

27.	Liaw A, Wiener M. Classification and regression by randomForest. R news. 2002;2(3):18-22.

28.	Chen T, He T, Benesty M, Khotilovich V, Tang Y, Cho H, et al. xgboost: Extreme Gradient Boosting. R package version 1.7.1.1 ed2022.

29.	Probst P, Wright MN, Boulesteix A-L. Hyperparameters and tuning strategies for random forest. WIREs Data Mining and Knowledge Discovery. 2019;9(3):e1301.

30.	Mayer M. kernelshap: Kernel SHAP. R package version 0.3.3 ed2023.

31.	Tierney N, Cook D, McBain M, Fay C. naniar: Data Structures, Summaries, and Visualisations for Missing Data. 0.6.1 ed2021.

32.	Sjoberg D. ggsankey: Sankey, Alluvial and Sankey Bump Plots. 0.0.99999 ed2022.

60.	Castellanos-Montiel MJ, Chaineau M, Durcan TM. The Neglected Genes of ALS: Cytoskeletal Dynamics Impact Synaptic Degeneration in ALS. Frontiers in Cellular Neuroscience. 2020;14.

67.	The UniProt C. UniProt: the universal protein knowledgebase in 2021. Nucleic Acids Res. 2021;49(D1):D480-D9.

68.	Chen H, Kankel MW, Su SC, Han SWS, Ofengeim D. Exploring the genetics and non-cell autonomous mechanisms underlying ALS/FTLD. Cell Death & Differentiation. 2018;25(4):648-62.

69.	Orlacchio A, Babalini C, Borreca A, Patrono C, Massa R, Basaran S, et al. SPATACSIN mutations cause autosomal recessive juvenile amyotrophic lateral sclerosis. Brain. 2010;133(2):591-8.

70.	Bunton-Stasyshyn RKA, Saccon RA, Fratta P, Fisher EMC. SOD1 Function and Its Implications for Amyotrophic Lateral Sclerosis Pathology:New and Renascent Themes. The Neuroscientist. 2015;21(5):519-29.

71.	Balendra R, Isaacs AM. C9orf72-mediated ALS and FTD: multiple pathways to disease. Nature Reviews Neurology. 2018;14(9):544-58.
